# Functional and structural neuroplasticity of motor system in hemiplegic cerebral palsy

**DOI:** 10.64898/2026.11.20.26368423

**Authors:** Sadra Shahdadian, Yanlong Song, Alireza Vaysi, Regan Christian, Madhan Bosemani, Fernando Acosta, Brooke Kimbrell, Warren Marks, Christos Papadelis

## Abstract

Hemiplegic cerebral palsy results from a unilateral prenatal or perinatal brain injury and causes motor impairments in the contralateral side of the body. Variable functional outcomes among children with hemiplegic cerebral palsy despite similar lesion extent suggest that developmental neuroplasticity and brain reorganization contribute to motor function. By using multimodal neuroimaging, we characterize the structural and functional reorganization of the motor system and examine its associations with lesion characteristics and upper-limb performance.

We examined 26 children with hemiplegic cerebral palsy and 26 typically developing controls using electromagnetic source imaging to map the primary motor cortex and characterize movement-related oscillations across physiologically relevant frequency bands, navigated transcranial magnetic stimulation to assess motor cortical excitability and lateralization, and functionally guided diffusion-weighted MRI to evaluate the structural integrity of descending motor and motor commissural fibers. We further assessed upper-limb function using Melbourne Assessment 2 and examined multimodal relationships using correlation, cross-validated latent-variable, and mediation analyses.

Children with hemiplegic cerebral palsy showed microstructural abnormalities in the descending motor and motor commissural fibers, characterized by lower fractional anisotropy and higher mean, axial, and radial diffusivity (all *P* < 0.05). The navigated transcranial magnetic stimulation identified ipsilateral corticospinal projections from the less affected hemisphere in 17 of 22 children (77%) with hemiplegic cerebral palsy, accompanied by reduced contralesional primary motor cortex lateralization (*P* < 0.05). The hemiplegic cerebral palsy group also exhibited hemisphere-specific abnormalities in movement-related oscillations, including reduced theta synchronization in the contralesional hemisphere and attenuated mu- and beta-band desynchronization in the ipsilesional hemisphere (all *P* < 0.05). Larger lesions were associated with poorer motor performance (*Rho* = −0.74 to −0.57, *P* < 0.05). Reduced descending motor fiber integrity was associated with impaired dexterity and attenuated mu- and beta-band desynchronization (*Rho* = −0.51 to 0.62, *P* < 0.05), while motor commissural fiber integrity was associated with corticospinal motor lateralization (*Rho* = −0.45 to 0.41, *P* < 0.05). A cross-validated multimodal structural-functional model predicted motor function (Q² = 0.42, *P* ≤ 0.001) more accurately than structural measures alone (Q² = 0.25). Beta-band desynchronization mediated the association between descending motor fiber integrity and motor performance (indirect effect = 0.12 SD, 95% CI 0.03–0.24).

Developmental motor neuroplasticity in hemiplegic cerebral palsy reflects an integrated structure-physiology-behavior process rather than the direct consequence of lesion burden alone. Multimodal biomarkers may improve prognostication and support individualized rehabilitation and neuromodulation.

## Introduction

Hemiplegic cerebral palsy (HCP) is a common subtype of cerebral palsy caused by unilateral perinatal injury that produces contralateral motor impairments.^1–6^ Affected children often experience spasticity, weakness, impaired motor control, and reduced manual dexterity, limiting independence and quality of life.^2,3,6^ Lesion characteristics influence the severity of these impairments, yet motor outcomes vary widely among children with comparable injuries. Function depends not only on the primary lesion but also on developmental neuroplasticity, the brain’s capacity to reorganize after early injury.^6–11^ The mechanisms of this reorganization remain poorly understood, limiting rehabilitation strategies tailored to individual neuroplastic potential.

This gap motivates identifying biomarkers of patient-specific motor reorganization.^7,12,13^ The developing brain can undergo substantial structural and functional adaptations after brain injury, including remodeling of corticospinal projections, cortical representations, and interhemispheric connections.^7–10^ Such adaptations may preserve motor function through alternative pathways, but they can also produce atypical network reorganization, such as mirror movements that impair movement selectivity and coordination.^14,15^ Characterizing the neural mechanisms of voluntary and stimulation-evoked motor control is essential to distinguish adaptive from maladaptive neuroplasticity in HCP and to guide personalized rehabilitation.

Structural MRI has clarified the anatomical basis of motor dysfunction in HCP, particularly the distinct consequences of periventricular (PV) and cortical-subcortical (CSC) lesions. Arising at different prenatal developmental stages, these lesions are associated with distinct patterns of structural damage, corticospinal reorganization, and motor impairment.^16,17^ PV lesions precede the corticospinal pathway maturation and therefore often leave greater capacity for adaptive reorganization,^18,19^ whereas CSC lesions more often disrupt established motor networks and entail greater upper-extremity dysfunction.^5–10,16,20^ Conventional MRI defines lesion type and extent without assessing motor-pathway integrity. Diffusion-weighted imaging (DWI) addresses this limitation and has revealed microstructural abnormalities in descending motor fibers (DMF) and motor commissural fibers (MCF) within the more affected (MA) hemisphere of children with HCP.^21–23^ Reduced DMF integrity is associated with poorer motor execution, dexterity, and functional outcomes,^20,21,24^ whereas MCF abnormalities, which reflect disrupted interhemispheric communication through the corpus callosum, relate to impaired bimanual coordination, reduced motor selectivity, and poorer rehabilitation response.^21,24–26^ Motor dysfunction in HCP thus reflects disruption across distributed pathways rather than injury confined to the primary lesion.

Structural imaging defines the anatomical substrate of motor dysfunction but cannot reveal how injury alters motor-system function. Functional neuroimaging, including magnetoencephalography (MEG) and high-density electroencephalography (HD-EEG), fills this gap by noninvasively measuring cortical movement-related responses (MRR).^27,28^ In cerebral palsy, these techniques have revealed altered sensorimotor oscillations and atypical cortical activation patterns.^29–34^ Disrupted transcallosal inhibition may compound these changes by promoting excessive bilateral cortical activation during unilateral movement, further reducing motor selectivity.^21,24,25^ Functional reorganization therefore extends beyond the primary lesion to distributed cortical networks, although its contribution to voluntary upper-extremity motor control remains poorly characterized.

While MEG and HD-EEG capture motor-network activity during voluntary movement, navigated transcranial magnetic stimulation (nTMS) probes corticospinal organization and cortical excitability directly.^35–37^ By eliciting motor-evoked potentials (MEPs) through focal stimulation of the primary motor cortex (M1), nTMS maps corticospinal projections and hemispheric motor organization.^38^ Excitability threshold is elevated in the ipsilesional motor cortex of children with HCP,^22^ and contralesional M1 stimulation can elicit MEPs in the affected limb, indicating the persistence of ipsilateral corticospinal projections.^39–41^ This pattern supports the hypothesis that early unilateral injury disrupts developmental pruning of bilateral corticospinal connections. These findings indicate that motor impairment arises from interactions among cortical activation, corticospinal organization, and interhemispheric communication.

Behavioral, structural, and functional measures inform neuroplasticity in HCP but are rarely integrated, leaving knowledge fragmented across interdependent levels. Here, we employ multimodal neuroimaging to characterize motor-system reorganization in children with HCP and to relate structural and functional markers of neuroplasticity to lesion characteristics and upper-extremity motor performance. We hypothesize that early unilateral brain injury initiates a cascade of structural and physiological adaptations, in which disruption of descending and commissural motor pathways constrains corticospinal organization and reshapes system-wide cortical motor activity, ultimately shaping motor outcome. We combined simultaneous MEG and HD-EEG recordings during a visuomotor task, nTMS assessment of corticospinal excitability and hemispheric organization, and diffusion tractography seeded from functionally localized motor cortex to quantify DMF and MCF microstructure. These measures were then related to standardized upper-extremity assessments using correlation, latent-variable, and mediation analyses.

## Materials and Methods

### Participants

We recruited 26 children with HCP (17 male; 10.8 ± 3.0 years) from the cerebral palsy clinic at Cook Children’s Medical Center, Fort Worth, Texas, and 26 typically developing (TD) children (14 male; 11.0 ± 3.1 years) from the local community. The groups did not differ in age (*P* = 0.82) or gender (*P* = 0.57).

Children with HCP were eligible if they had: (1) diagnosis confirmed by a pediatric neurologist, developmental specialist, or neonatologist; (2) Gross Motor Function Classification System level I or II; and (3) cognitive ability to follow age-appropriate instructions. TD children were screened to exclude neurological, developmental, or sensory disorders. Exclusion criteria for both groups were: (1) genetic syndromes; (2) prior brain surgery or major traumatic brain injury; (3) inability to remain still during recordings; and (4) contraindications to MEG or MRI. The Cook Children’s Health Care System Institutional Review Board approved all procedures (2019-068; PI: C. Papadelis). Written parental consent and child assent, as appropriate, were obtained from all participants.

### Movement quality

Upper limb motor function was assessed using the Melbourne Assessment 2 (MA2; Fig. 1A).^42^ Participants completed standardized unilateral upper-extremity tasks evaluating range of motion (ROM), accuracy, dexterity, and fluency. Assessments were video-recorded and independently scored offline by two raters (E.A. and R.C.) according to MA2 administration and scoring guidelines. Standardized scores were used in subsequent analyses.

**Figure 1.**
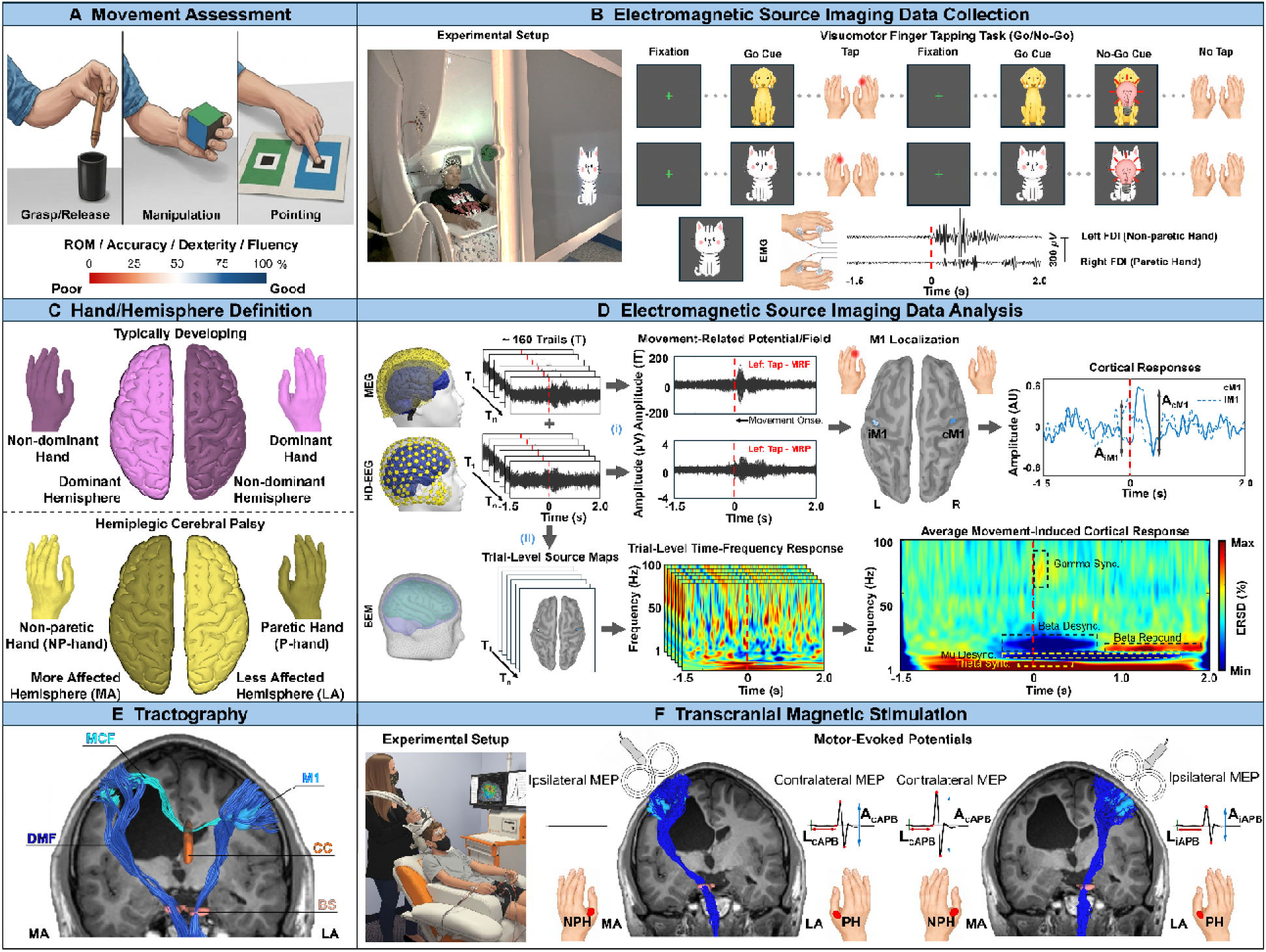
Multimodal assessment of motor-system organization and analysis workflow. **(A)** Upper-limb motor function was assessed using the Melbourne Assessment 2 test, comprising range of motion (ROM), accuracy, dexterity, and fluency. Representative tasks include grasp/release, manipulation, and pointing. **(B)** Simultaneous magnetoencephalography (MEG) and high-density EEG (HD-EEG) were recorded during a Go/NoGo visuomotor task. Dog and cat visual cue (Go) instructed right- and left-index-finger tapping, respectively; a subsequent lightbulb cue (NoGo) instructed response inhibition. Bilateral first dorsal interosseous (FDI) electromyography (EMG) identified movement onset (time zero). **(C)** Hand and hemisphere definitions for typically developing (TD) children and children with hemiplegi cerebral palsy (HCP). Dominant and nondominant hemispheres were defined contralateral to the corresponding hands. In HCP, the more affected (MA) hemisphere was contralateral to the paretic hand (P-hand), and the less affected (LA) hemisphere was contralateral to the nonparetic hand (NP-hand). **(D)** Electromagnetic source imaging followed two complementary analysis pathways using individual MRI-derived boundary element models (BEM): (i) Movement-onset-aligned trials were averaged to obtain movement-related fields (MRF) and potentials (MRP), and source reconstruction localized bilateral primary motor cortices (M1). Response amplitudes in contralateral (cM1) and ipsilateral (iM1) M1 were used to calculate movement-related lateralization. (ii) Single-trial source time series underwent time– frequency decomposition, averaging and baseline normalization to quantify movement-induced oscillatory responses. Highlighted components comprise theta synchronization, mu and bet desynchronization, post-movement beta rebound and gamma synchronization. **(E)** Diffusion tractography combined functionally defined M1 regions with anatomical brainstem (BS) and corpus callosum (CC) regions to reconstruct descending motor fibers (DMF) and motor commissural fibers (MCF), respectively. Diffusion metrics were extracted to characterize tract microstructure. **(F)** Navigated transcranial magnetic stimulation (nTMS) mapped motor output from each hemisphere while motor-evoked potentials (MEPs) were recorded bilaterally from the abductor pollicis brevis (APB). MEP amplitude and latency characterized corticospinal responses. Bilateral amplitudes were used to calculate stimulation-evoked lateralization. The illustrated example shows contralateral responses following MA-hemisphere stimulation and bilateral responses following LA-hemisphere stimulation. A: amplitude; L: latency; c: contralateral; i: ipsilateral; AU: arbitrary units; ERSD: event-related synchronization/desynchronization; NPH/PH: nonparetic/paretic hand.

### MEG and HD-EEG acquisition

Simultaneous MEG and HD-EEG were recorded during a visuomotor task (Fig. 1B) in a single-layer magnetically shielded room in the Radiology suite at Cook Children’s Medical Center. MEG was acquired using a whole-head 306-channel VectorView® system (Elekta Neuromag, Helsinki, Finland; 102 magnetometers and 204 planar gradiometers). HD-EEG was recorded using an EGI Geodesic system (Electrical Geodesics, Eugene, OR) with a 128-or 256-channel saline-based net selected according to head size. MEG and HD-EEG data were sampled at 1 kHz, and electrode impedances were maintained below 50 kΩ.

Electrode locations were digitized relative to the head using the GeoScan Sensor Digitization system (Magstim EGI, USA). Five head-position indicator coils were placed on the scalp for MEG head-position tracking. Before each recording, the nasion, bilateral preauricular points, and ∼500 scalp-reference points were digitized with a Polhemus Fastrak system.

Peripheral signals were also recorded including: (1) the bilateral surface electromyography (EMG) from the first dorsal interosseous muscles to identify motor-response onset (Fig. 1B); (2) vertical electrooculography (EOG) from bipolar electrodes placed above and below the right eye to identify ocular artifacts; and (3) electrocardiography (ECG) from electrodes placed over the upper chest to identify cardiac artifacts for subsequent removal during preprocessing.

### MRI and diffusion-weighted imaging (DWI) acquisition

Structural T1-weighted MRI and DWI were acquired on a 3T Siemens Skyra MRI scanner using a 10-channel head coil. T1-weighted images were obtained with a 3D magnetization-prepared rapid-acquisition gradient-echo (MPRAGE) sequence (echo time□=□3.22 ms, inversion time□=□450 ms, repetition time□=□8.21 ms, flip angle□=□15 degrees, field of view□=□24 cm, matrix size□=□256□×□256, 184 slices, and resolution□=□1.0□×□1.0□×□1.0 mm³). These images were used for cortical surface reconstruction and forward modeling for MEG/HD-EEG source estimation.

DWI was acquired using a multi-shell diffusion protocol with b-values of 1,000 and 2,000 s/mm², with 30 and 60 diffusion-encoding directions, respectively. Images had 2.05 mm in-plane resolution and 2 mm slice thickness.

### Radiological findings

A pediatric neuroradiologist (M.B.) and pediatric neurologist (W.M.) reviewed MRI scans to classify lesion type, laterality, and involvement of cortical gray matter, periventricular white matter, basal ganglia, thalamus, and corpus callosum. In children with HCP, the hemisphere with the most pronounced structural abnormality was designated the MA hemisphere and the contralateral hemisphere the less affected (LA) hemisphere (Fig. 1C). The MA hemisphere was contralateral to the paretic hand (P-hand) in all participants. Participant demographic and clinical characteristics are provided in Table 1.

**Table 1.**
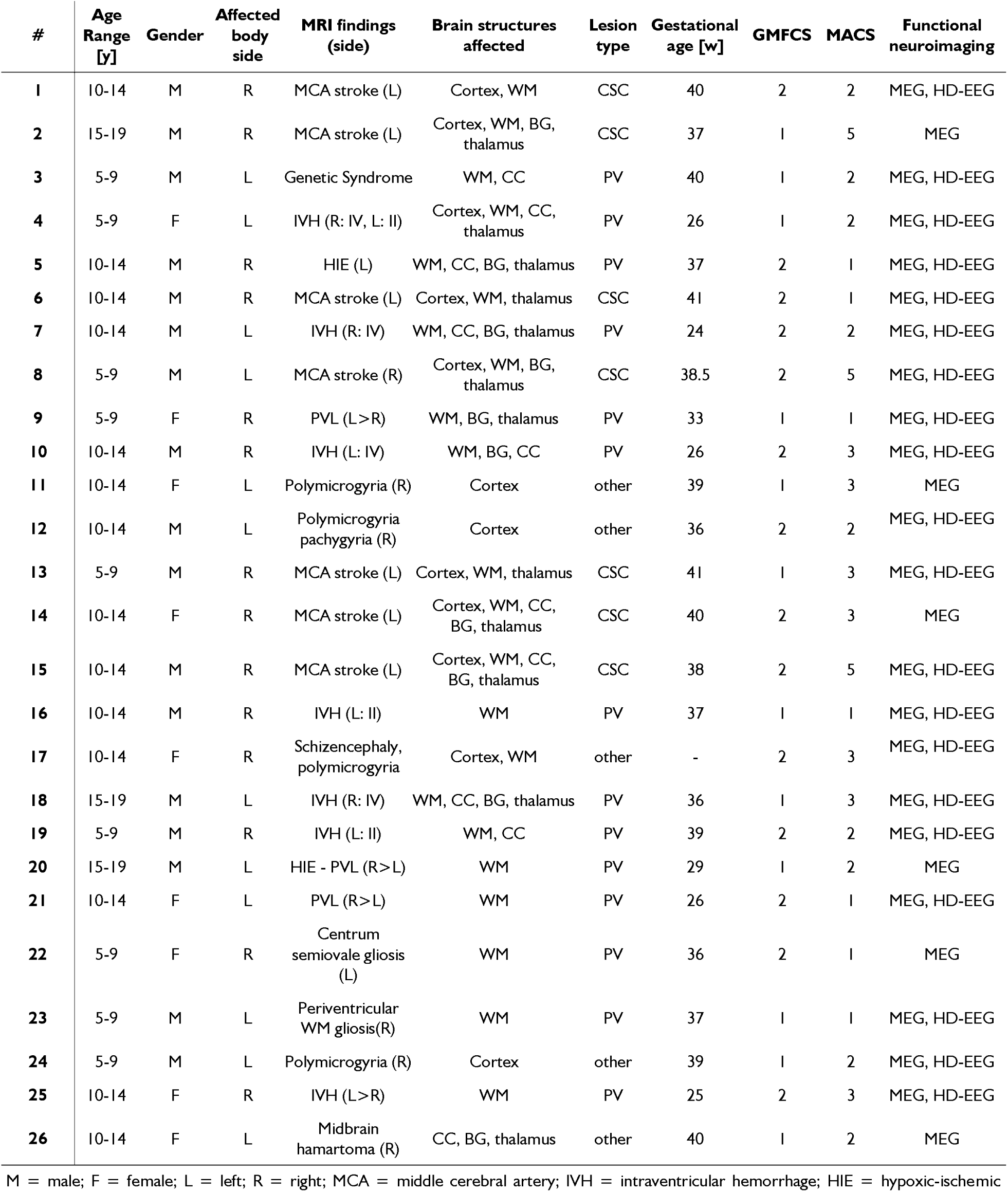
demographics and clinical characteristics of hemiplegic cerebral palsy participants.

| **#** | **Age Range [y]** | **Gender** | **Affected body side** | **MRI findings (side)** | **Brain structures affected** | **Lesion type** | **Gestational age [w]** | **GMFCS** | **MACS** | **Functional neuroimaging** |
|---|---|---|---|---|---|---|---|---|---|---|
| **1** | 10-14 | M | R | MCA stroke (L) | Cortex, WM | CSC | 40 | 2 | 2 | MEG, HD-EEG |
| **2** | 15-19 | M | R | MCA stroke (L) | Cortex, WM, BG, thalamus | CSC | 37 | 1 | 5 | MEG |
| **3** | 5-9 | M | L | Genetic Syndrome | WM, CC | PV | 40 | 1 | 2 | MEG, HD-EEG |
| **4** | 5-9 | F | L | IVH (R: IV, L: II) | Cortex, WM, CC, thalamus | PV | 26 | 1 | 2 | MEG, HD-EEG |
| **5** | 10-14 | M | R | HIE (L) | WM, CC, BG, thalamus | PV | 37 | 2 | 1 | MEG, HD-EEG |
| **6** | 10-14 | M | R | MCA stroke (L) | Cortex, WM, thalamus | CSC | 41 | 2 | 1 | MEG, HD-EEG |
| **7** | 10-14 | M | L | IVH (R: IV) | WM, CC, BG, thalamus | PV | 24 | 2 | 2 | MEG, HD-EEG |
| **8** | 5-9 | M | L | MCA stroke (R) | Cortex, WM, BG, thalamus | CSC | 38.5 | 2 | 5 | MEG, HD-EEG |
| **9** | 5-9 | F | R | PVL (L>R) | WM, BG, thalamus | PV | 33 | 1 | 1 | MEG, HD-EEG |
| **10** | 10-14 | M | R | IVH (L: IV) | WM, BG, CC | PV | 26 | 2 | 3 | MEG, HD-EEG |
| **11** | 10-14 | F | L | Polymicrogyria (R) | Cortex | other | 39 | 1 | 3 | MEG |
| **12** | 10-14 | M | L | Polymicrogyria pachygyria (R) | Cortex | other | 36 | 2 | 2 | MEG, HD-EEG |
| **13** | 5-9 | M | R | MCA stroke (L) | Cortex, WM, thalamus | CSC | 41 | 1 | 3 | MEG, HD-EEG |
| **14** | 10-14 | F | R | MCA stroke (L) | Cortex, WM, CC, BG, thalamus | CSC | 40 | 2 | 3 | MEG |
| **15** | 10-14 | M | R | MCA stroke (L) | Cortex, WM, CC, BG, thalamus | CSC | 38 | 2 | 5 | MEG, HD-EEG |
| **16** | 10-14 | M | R | IVH (L: II) | WM | PV | 37 | 1 | 1 | MEG, HD-EEG |
| **17** | 10-14 | F | R | Schizencephaly, polymicrogyria | Cortex, WM | other | - | 2 | 3 | MEG, HD-EEG |
| **18** | 15-19 | M | L | IVH (R: IV) | WM, CC, BG, thalamus | PV | 36 | 1 | 3 | MEG, HD-EEG |
| **19** | 5-9 | M | R | IVH (L: II) | WM, CC | PV | 39 | 2 | 2 | MEG, HD-EEG |
| **20** | 15-19 | M | L | HIE - PVL (R>L) | WM | PV | 29 | 1 | 2 | MEG |
| **21** | 10-14 | F | L | PVL (R>L) | WM | PV | 26 | 2 | 1 | MEG, HD-EEG |
| **22** | 5-9 | F | R | Centrum semiovale gliosis (L) | WM | PV | 36 | 2 | 1 | MEG |
| **23** | 5-9 | M | L | Periventricular WM gliosis(R) | WM | PV | 37 | 1 | 1 | MEG, HD-EEG |
| **24** | 5-9 | M | L | Polymicrogyria (R) | Cortex | other | 39 | 1 | 2 | MEG, HD-EEG |
| **25** | 10-14 | F | R | IVH (L>R) | WM | PV | 25 | 2 | 3 | MEG, HD-EEG |
| **26** | 10-14 | F | L | Midbrain hamartoma (R) | CC, BG, thalamus | other | 40 | 1 | 2 | MEG |

M = male; F = female; L = left; R = right; MCA = middle cerebral artery; IVH = intraventricular hemorrhage; HIE = hypoxic-ischemic [illegible]

For TD participants, the hemisphere contralateral to the dominant hand (D-hand) was designated the dominant hemisphere (D-hemi).

### Lesion and ventricular dilation volume

Lesion and ventricular dilatation volumes were quantified from T1-weighted MRI.^5^ Lesion volume included cortical gray-matter loss and ex-vacuo ventricular dilatation attributable to white-matter volume loss. To account for normal ventricular volume, we subtracted the mean ventricular volume of the TD group from the combined lesion and enlarged ventricular volume in the MA hemisphere.

### Visuomotor task

Participants completed a Go/NoGo visuomotor task during simultaneous MEG and HD-EEG recording to elicit cortical activity related to movement preparation, execution, and response inhibition (Fig. 1B). Visual stimuli were presented centrally on a back-projection screen placed ∼150 cm in front of the participant. Stimulus presentation and behavioral responses were controlled with Stim2 software (Compumedics Neuroscan, Charlotte, NC, USA) with event triggers synchronized to the MEG and HD-EEG systems for subsequent epoching and analysis.

Each trial began with a 500-ms fixation cross followed by a 1,200-ms dog or cat Go cue (Fig. 1B). Participants were instructed to tap the right index finger in response to the dog and left index finger in response to the cat. On pseudorandomly selected trials, a cartoon light bulb appeared 230–440 ms after cue onset (serving as the NoGo cue) signaling participants to withhold the planned response. Each run included 120 trials (60 dog and 60 cat cues), including 30 NoGo trials equally distributed across cue types. Participants completed four 8-minute runs.

### Preprocessing of MEG/HD-EEG data

Raw MEG data were processed using *MaxFilter* (version 2.2.10, MEGIN, Finland) with temporal signal-space separation to suppress noise and correct for head movement. MEG and HD-EEG data were subsequently spatially aligned and temporally synchronized using an in-house algorithm.^43^

MEG and HD-EEG channels with excessive artifacts were identified by visual inspection and removed. Data were baseline-corrected, band-pass filtered (1-100 Hz), and notch-filtered at 60 Hz using *Brainstorm*.^44^ MEG artifacts related to cardiac or ocular activity were identified from ECG and EOG recordings and removed using signal-space projection.^33^ HD-EEG artifacts arising from cardiac activity, eye movements, blinks, and muscle activity were removed using independent component analysis.^34^ For datasets with prominent muscle artifacts, components most strongly associated with contaminated channels were removed. Remaining noisy data segments and their corresponding trials were excluded after visual inspection.

Bilateral first dorsal interosseous EMG signals were band-pass filtered (30–200 Hz) and notch-filtered at 60, 120, and 180 Hz. EMG onset following Go cues was manually identified and used for epoching.

### Electromagnetic source localization

Continuous MEG/HD-EEG data were segmented into 3,500 ms epochs time-locked to EMG-defined movement onset (1,500 prior and 2,000 ms after movement). Trials were averaged within participant to derive movement-related fields and potentials, and channels with excessive premovement activity were excluded (Fig 1D).

We estimated cortical source activity using dynamic Statistical Parametric Mapping (dSPM).^45,46^ Individual T1-weighted MRI scans were used to generate cortical, skull, and scalp surfaces. MEG/HD-EEG data were coregistered to each participant’s anatomy using digitized head points, and forward models were computed with boundary element modeling (BEM) in *OpenMEEG*.^47^ Noise covariance matrices were estimated and used to solve the inverse model across ∼15,000 cortical vertices using combined gradiometer, magnetometer, and HD-EEG data, with dipole orientations normal to the cortical surface.

Primary motor cortex (M1) was identified as the peak movement-related activity within Brodmann area 4 at 120-150 ms^48^ after movement onset. The peak and adjacent vertices were expanded to define an ∼3 cm^2^ M1 region of interest for diffusion tractography (Fig. 1E). In two participants with severe cortical lesions, movement-related peaks for the P-hand did not align with the anatomical atlas; therefore, the M1 region was defined from the peak activity in the MA hemisphere.

### Time–frequency analysis of movement-induced response

Single-trial time series extracted from virtual M1 vertex were decomposed into time-frequency representations using complex Morlet wavelets (central frequency of 1 Hz and a cycle count of three cycles). To quantify non-phase-locked, movement-induced activity, time-frequency maps were averaged across trials for each participant. Event-related synchronization (ERS) and desynchronization (ERD) were calculated as power changes relative to the premovement baseline (−1,500 to −1,000 ms).^49^

### Spectrotemporal components of movement-induced responses

Time-frequency maps were examined across five movement-related spectrotemporal components implicated in motor initiation, execution, and inhibition: theta band synchronization (ERS_theta_; 4– 8 Hz), mu desynchronization (ERD_mu_; 8–12 Hz), beta desynchronization (ERD_beta_; 13–30 Hz), post-movement beta rebound (PMBR_beta_; 16–30 Hz), and gamma synchronization or burst (ERS_gamma_; 30–80 Hz). For each participant, baseline-normalized power was averaged within each time-frequency window and compared between groups.

### Navigated Transcranial Magnetic Stimulation

Individual T1-weighted MRI scans were reconstructed into three-dimensional head models and coregistered to each participant for navigated TMS (nTMS; Nexstim NBS 5.1, Helsinki, Finland; Fig. 1F). Bilateral EMG was recorded from the abductor pollicis brevis (APB), with reference electrodes placed on the tendon and a ground electrode on the hand dorsum. Signals were sampled at 3 kHz (10-500 Hz bandwidth, 0.3μV resolution).

Single-pulse nTMS was delivered using a figure-of-eight coil and frameless stereotactic neuronavigation. The precentral gyrus was mapped in each hemisphere to identify the APB motor hotspot. Resting motor threshold (rMT) was defined as the minimum intensity that elicited MEP of at least 50 μV in 10 of 20 trials. Motor mapping was then performed at 110% rMT with the coil-oriented perpendicular to the cortical gyrus in a dorsal-rostral direction to optimize corticospinal tract activation. Both hemispheres were mapped while recording bilateral APB EMG activity.

### Motor lateralization index

Lateralization indices (LI) were calculated to quantify interhemispheric balance during voluntary movement and nTMS-evoked responses. To estimate the voluntary movement-related cortical LI (LI_MRR_), we extracted baseline-normalized movement-related response amplitudes from bilateral MRR-defined primary motor cortices (M1) during unilateral finger movements (Fig. 1D). To estimate the nTMS-evoked responses (LI_TMS_), we extracted the MEP amplitudes from bilateral APB muscles during motor mapping. Then, we calculated the LI as:

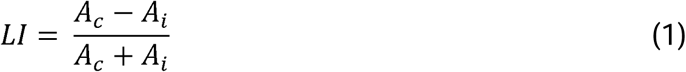

For LI_MRR_, *A_c_* and *A_i_* represent cortical activity (baseline-normalized MRR amplitude) in the contralateral and ipsilateral M1, respectively. For LI_TMS_, *A_c_* and *A_i_*represent MEP amplitudes in the contralateral and ipsilateral APB, respectively.^50^ The LI_TMS_ was calculated for each stimulation site and averaged within hemisphere for each participant (Fig. 1F). *LI* values range from –1, indicating ipsilateral activity, to +1 indicating exclusively contralateral activity. Together, LI_MRR_ and LI_TMS_ characterize interhemispheric balance during voluntary and stimulation-evoked motor responses.

LIs were also calculated for movement-related spectrotemporal components. Because these measures may have positive or negative values, a modified LI formula was used as described in the Supplementary materials.

### ROI-based tractography

DWI data were preprocessed and analyzed in *DSI-Studio* (version October 10, 2024). Preprocessing included visual quality control, motion correction, and white-matter mask generation. Fiber tracking used generalized Q-sampling imaging, which resolves complex fiber configurations, including crossing fibers.^51^ Tracking parameters included a 60° angular threshold, 0.5-1.5 voxels step sizes, and 30-200 mm tract lengths. Topology-informed pruning was applied over 16 iterations to remove false-positive correlations from an initial set of 5,000 tracts. ^52^

Functionally defined M1 regions were imported into *DSI studio* for subject-specific tractography (Fig. 1E). DMFs were reconstructed using M1 and the ventral brainstem corticospinal tract as regions of interest. MCF were identified by seeding from unilateral M1 with the corpus callosum (CC) as an additional region of interest; tracts were then restricted to the ipsilateral hemisphere. Fractional anisotropy (FA), mean diffusivity (MD), axial diffusivity (AD), and radial diffusivity (RD) were calculated for each reconstructed tract to characterize white-matter microstructural integrity. The same tractography procedure was performed using the nTMS-defined ROIs.

### Statistical analysis

Within-group comparisons across hands and hemispheres used *Wilcoxon* signed rank tests. These analyses included MA2 outcomes; response time and inhibition accuracy; movement-related and TMS-evoked LI (LI_MRR_ and LI_TMS_); latency and resting-state motor threshold; and diffusion metrics (FA, MD, AD, and RD).

Between-group comparisons used *Mann-Whitney* U tests with false-discovery-rate correction. We compared the TD nondominant hand/hemisphere with the HCP paretic hand/more affected hemisphere and the TD dominant hand/hemisphere with the HCP nonparetic hand/less affected hemisphere. Statistical significance was set at 0.05. Effect sizes are reported as the absolute rank-biserial correlation (0–1).

### Inter-modality correlation

Spearman correlations assessed associations among behavioral, functional, and structural measures, focusing on functional-behavioral, structural-behavioral, and structural-functional relationships. Analyses were conducted across all hands and hemispheres in both groups and separately within the P-hand/MA hemisphere of the HCP group. Statistical significance was set at 0.05. The analyses were performed in R® version 4.5.0.

### Latent structural-functional integrity score

We used partial least-squares (PLS) regression to test whether a multivariate structural-functional integrity score predicted overall motor outcome. PLS was selected because diffusion metrics within each tract were highly collinear (correlations up to 0.9 among FA, MD, AD, and RD). The model integrated diffusion metrics from DMF and MCF with movement-related MEG/EEG oscillatory measures and their LIs. Model performance was evaluated using leave-one-out cross-validation, with the number of PLS components selected by nested cross-validation within each training fold. Statistical significance was assessed using 1,000 permutation iterations. Separate PLS models incorporating DMF or MCF measures alone were used to determine whether each tract contributed independent predictive information.

### Mediation analysis

To test whether structural alterations influence motor function through cortical motor physiology, we examined whether ERD_beta_ mediated the association between DMF FA/MD/AD/RD and the logit-transformed composite MA2 score. Sequential regression with 5,000 bootstrap resamples tested the hypothesized intermediate role of cortical physiology in the structure–physiology– behavior relationship. ERD_beta_ was selected a priori because it is closely associated with movement-related sensorimotor cortical engagement and motor performance.^28,53^ PLS and mediation analyses were performed in Python® version 3.14.

## Results

### Participant characteristics and lesion distribution

Structural MRI showed predominantly unilateral lesions in children with HCP. Based on neuroradiological classification, seven participants had cortical-subcortical (CSC) lesions (6 male, 11.0 ± 3.2 years), 14 had periventricular (PV) lesions (10.5 ± 3.4 years), and five had other lesion types (11.0 ± 1.6 years). Detailed lesion characteristics are reported in Table 1.

### Upper-extremity motor performance

Children with HCP exhibited poorer P-hand performance than TD controls across all MA2 domains (Fig. 2A): range of motion (*P* < 0.001, r = 0.69), accuracy (*P* < 0.01, r = 0.41), dexterity (*P* < 0.001, r = 0.79), and fluency (*P* < 0.001, r = 0.65). Deficits were more pronounced among children with CSC lesions, who showed poorer motor performance than those with PV lesions, particularly with dexterity (*P* < 0.001, r = 0.91) (Fig. 2B). In contrast, the non-paretic hand (NP-hand) performance was largely preserved.

**Figure 2.**
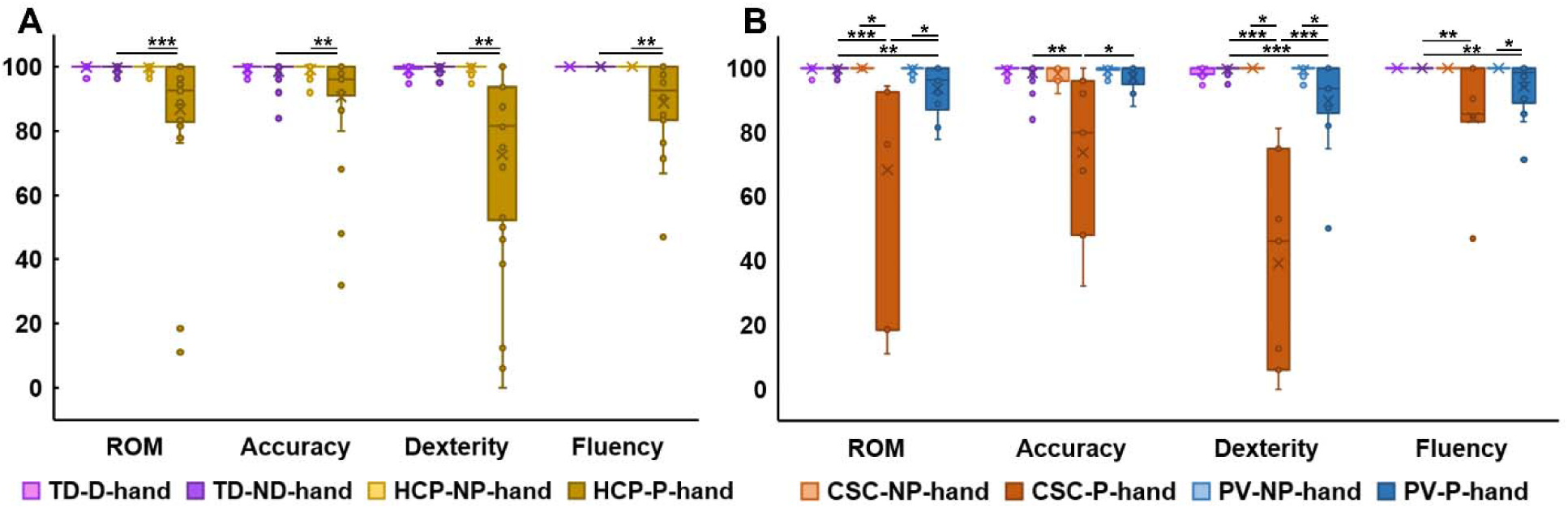
Upper-limb motor performance in hemiplegic cerebral palsy. Box-and-whisker plots show Melbourne Assessment 2 scores (%) for range of motion (ROM), accuracy, dexterity and fluency; higher scores indicate better performance. **(A)** Scores for the dominant (D-hand) and nondominant (ND-hand) hands of typically developing children (TD; n = 26) and the nonparetic (NP-hand) and paretic (P-hand) hands of children with hemiplegic cerebral palsy (HCP; n = 26). **(B)** Scores stratified by cortical– subcortical (CSC; n = 7) and periventricular (PV; n = 14) lesion types, with TD scores shown for comparison. Boxes represent the interquartile range, horizontal lines indicate medians, crosses denote means and circles represent individual observations. Within-participant comparisons used Wilcoxon signed-rank tests, and between-group comparisons used Mann–Whitney U tests with false-discovery-rate correction. Horizontal brackets indicate significant comparisons: ∗ *P* < 0.05, ∗∗ < 0.01, ∗∗∗ < 0.001.

### Response time and inhibition accuracy

During the visuomotor task, response times were longer for the P-hand than the NP-hand in children with HCP (*P* < 0.01, r = 0.67) and longer than for the TD nondominant hand (*P* < 0.001, r = 0.68; Fig. 3A). The HCP NP-hand was also slower than the TD dominant hand (*P* < 0.05, r = 0.39). Inhibition accuracy was higher (*P* < 0.05, r = 0.65) for the P-hand than the NP-hand (Fig. 3B).

**Figure 3.**
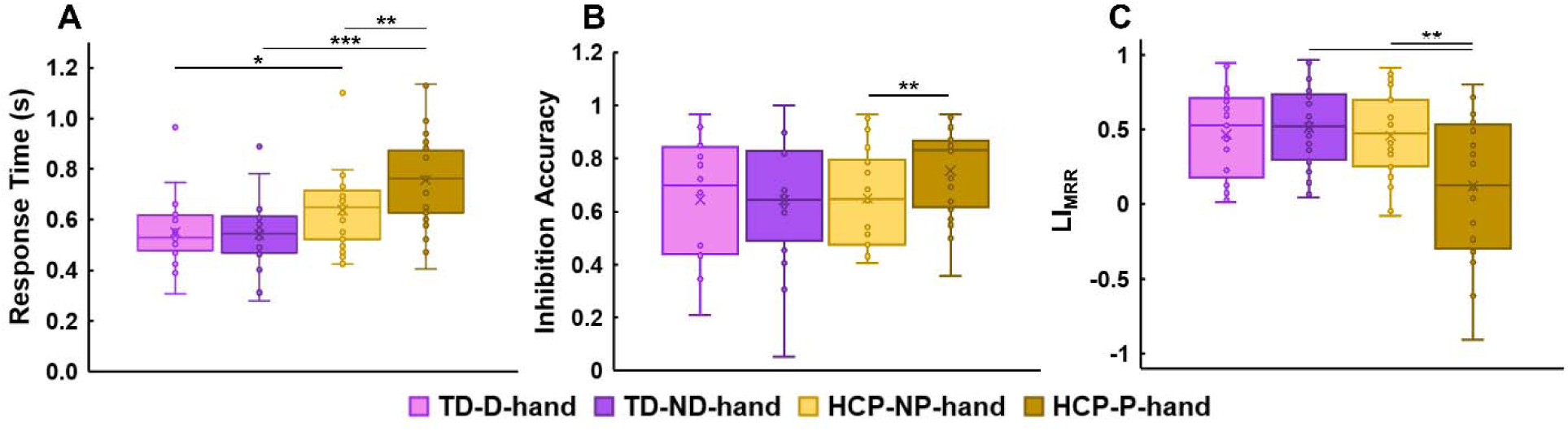
Visuomotor task performance and movement-related cortical lateralization. Box-and-whisker plots show **(A)** response time, **(B)** inhibition accuracy and **(C)** movement-related response lateralization index (LI_MRR_) during the Go/NoGo task. Data are shown for the dominant (D-hand) and nondominant (ND-hand) hands of typically developing children (TD; n = 26) and the nonparetic (NP-hand; n = 26) and paretic (P-hand; n = 23) hands of children with hemiplegic cerebral palsy (HCP). Response time reflects the interval between the Go cue and movement onset, whereas inhibition accuracy represents the proportion of NoGo trials in which the response was successfully withheld. LI_MR_ quantifies the balance of movement-related response amplitudes between contralateral and ipsilateral primary motor cortices, ranging from −1 (exclusively ipsilateral) to +1 (exclusively contralateral), with zero indicating equal bilateral amplitudes. Boxes represent the interquartile range, horizontal lines indicate medians, crosses denote means and circles represent individual observations. Within-participant comparisons used Wilcoxon signed-rank tests, and between-group comparisons used Mann–Whitney U tests with false-discovery-rate correction. Horizontal brackets indicate significant comparisons: ∗ *P* < 0.05, ∗∗ < 0.01, ∗∗∗ < 0.001.

### Movement-related lateralization index

Movement-related cortical activity indicated altered motor-cortical lateralization in HCP (Fig. 3C). During P-hand movement, the movement-related response lateralization index was lower than the TD group (*P* < 0.01, r = 0.50) and the HCP NP-hand (*P* < 0.05, r = 0.62), consistent with increased ipsilateral and/or reduced contralateral M1 activity.

### Movement-induced oscillatory activity

Time-frequency analysis demonstrated altered motor-related oscillatory dynamics in children with HCP (Fig. 4). During NP-hand movement, ERS_theta_ was reduced compared to controls (*P* < 0.05, Fig. 4A). During P-hand movement, children with HCP showed reduced ERD_mu_ in both contralateral and ipsilateral M1 compared with TD (*P* < 0.05; Fig. 4B and Fig. 4D). ERD_beta_ wa also reduced in contralateral M1 during P-hand movement and bilaterally during NP-hand movement (*P* < 0.05; Fig. 4B, Fig. 4C, and Fig. 4D). Supplementary Fig. 1 presents statistically significant differences among comparisons.

**Figure 4.**
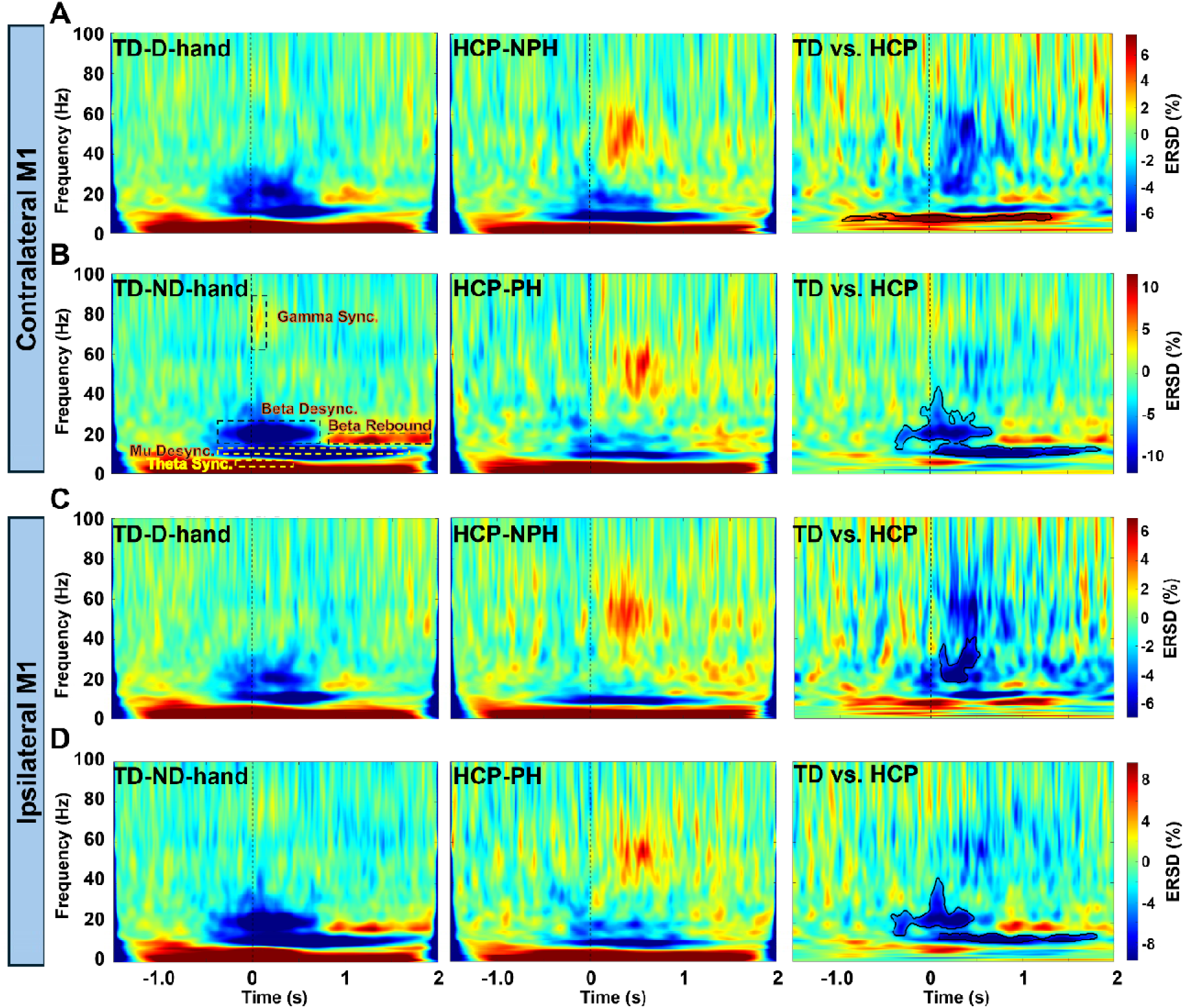
Movement-induced oscillatory activity in bilateral primary motor cortices. Grand-average time–frequency maps show cortical power changes during finger tapping in typically developing (TD) children and children with hemiplegic cerebral palsy (HCP). Columns display TD responses, HCP responses and between-group differences (TD − HCP), respectively. **(A, B)** Contralateral primary motor cortex (M1) responses during movement of the TD dominant hand (D-hand) versus HCP nonparetic hand (NPH), and the TD nondominant hand (ND-hand) versus HCP paretic hand (PH), respectively. **(C, D)** Corresponding responses in ipsilateral M1. Sample sizes were n = 26 for each TD hand and the HCP nonparetic hand, and n = 23 for the HCP paretic hand. Power was normalized to the premovement baseline (−1.5 to −1.0 s) and expressed as percentage change. In the group-average maps, warm color indicate event-related synchronization (ERS) and cool colors indicate desynchronization (ERD); in the difference maps, colors represent signed TD − HCP differences. Vertical dashed lines mark EMG-defined onset (0 s). Dashed rectangles in panel B identify the spectrotemporal windows for theta synchronization, mu and beta desynchronization, post-movement beta rebound and gamma synchronization. Black contours in the difference maps delineate significant between-group clusters identified using cluster-based permutation testing ( < 0.05). ERSD, event-related synchronization/desynchronization; EMG, electromyography.

### Spectrotemporal motor lateralization

Spectrotemporal lateralization differed in children with HCP (Supplementary Fig. 2). ERD_beta_ lateralization was greater during NP-hand movement (*P* < 0.05, r = 0.39), whereas ERS_gamma_ lateralization was reduced during P-hand movement (*P* < 0.05, r = 0.35).

### nTMS

No MEP was elicited by stimulation of either hemisphere in four children with HCP; an additional nine children had no MEP following the MA hemisphere stimulation. Among the remaining participants, nTMS demonstrated atypical corticospinal organization (Fig. 5). Stimulation of the LA hemisphere elicited bilateral MEPs in 17 of 22 children with HCP. Ipsilateral MEP latency was longer than contralateral MEP latency (*P* < 0.05, r = 0.49; Fig. 5A). The rMT for the contralateral APB responses was higher in the MA hemisphere (*P* < 0.05, r = 0.91; Fig. 5B). Bilateral responses after stimulation of the LA hemisphere resulted in LI_TMS_ < 1, whereas no ipsilateral MEPs were observed in TD children or following the MA hemisphere stimulation in children with HCP (*P* < 0.001, r > 0.77; Fig. 5C).

**Figure 5.**
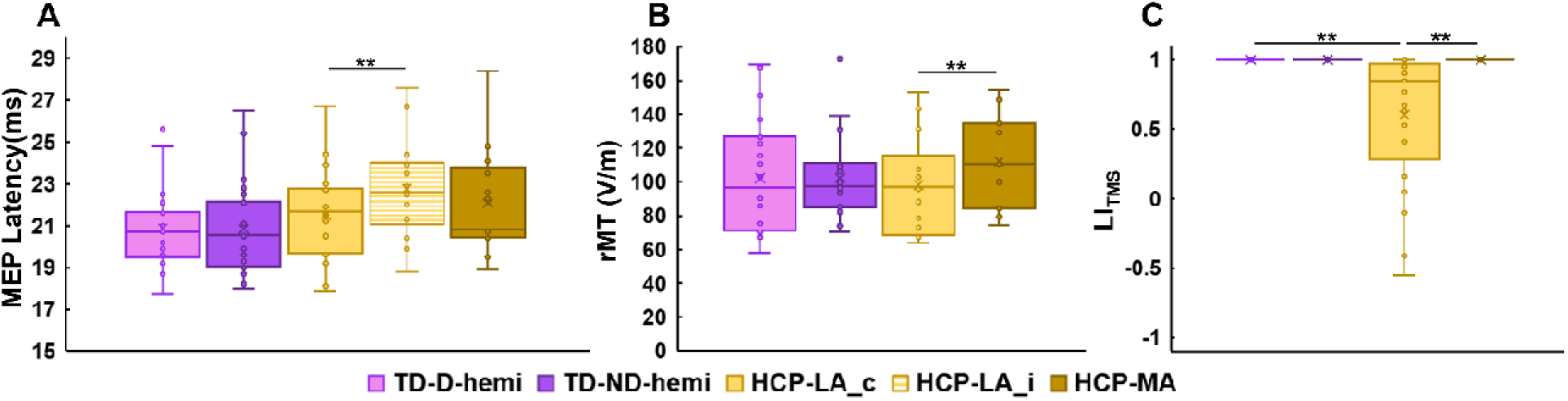
Corticospinal excitability and motor-output lateralization assessed using navigated transcranial magnetic stimulation (nTMS). Box-and-whisker plots show **(A)** motor-evoked potential (MEP) latency, **(B)** resting motor threshold (rMT) and **(C)** stimulation-evoked lateralization index (LI_TMS_). Data are shown for the dominant (D-hemi) and nondominant (ND-hemi) hemispheres of typically developing children (TD; n = 24) and the less affected (LA; n = 22) and more affected (MA; n = 13) hemispheres of children with hemiplegic cerebral palsy (HCP). **(A)** MEP latencies were recorded from the abductor pollicis brevis contralateral to the stimulated hemisphere. Ipsilateral responses following LA-hemisphere stimulation are shown separately as striped boxes (HCP-LA_i; n = 17); HCP-LA_c denote contralateral responses. **(B)** Resting motor threshold is expressed as electric field strength (V/m). **(C)** LI_TMS_ quantifies the relative amplitudes of contralateral and ipsilateral MEPs, ranging from −1 (exclusively ipsilateral) to +1 (exclusively contralateral), with zero indicating equal bilateral amplitudes. Boxes represent the interquartile range, horizontal lines indicate medians, crosses denote means and circles represent individual observations. Within-participant comparisons used Wilcoxon signed-rank tests, and between-group comparisons used Mann–Whitney U tests with false-discovery-rate correction. Horizontal brackets indicate significant comparisons: ∗∗ *P* < 0.01.

### Motor-pathway microstructure

Diffusion tractography demonstrated reduced DMF microstructural integrity in the MA hemisphere (Fig. 6A). Relative to the TD group and the LA hemisphere of the HCP group, the DMF of the MA hemisphere showed lower FA (*P* < 0.01, r > 0.71) and higher MD (*P* < 0.001, r > 0.85), AD (*P* < 0.01, r > 0.50), and RD (*P* < 0.001, r > 0.79). Within the MA hemisphere, the CSC group had higher AD than the PV group (*P* < 0.05, r = 0.83; Supplementary Fig. 3).

**Figure 6.**
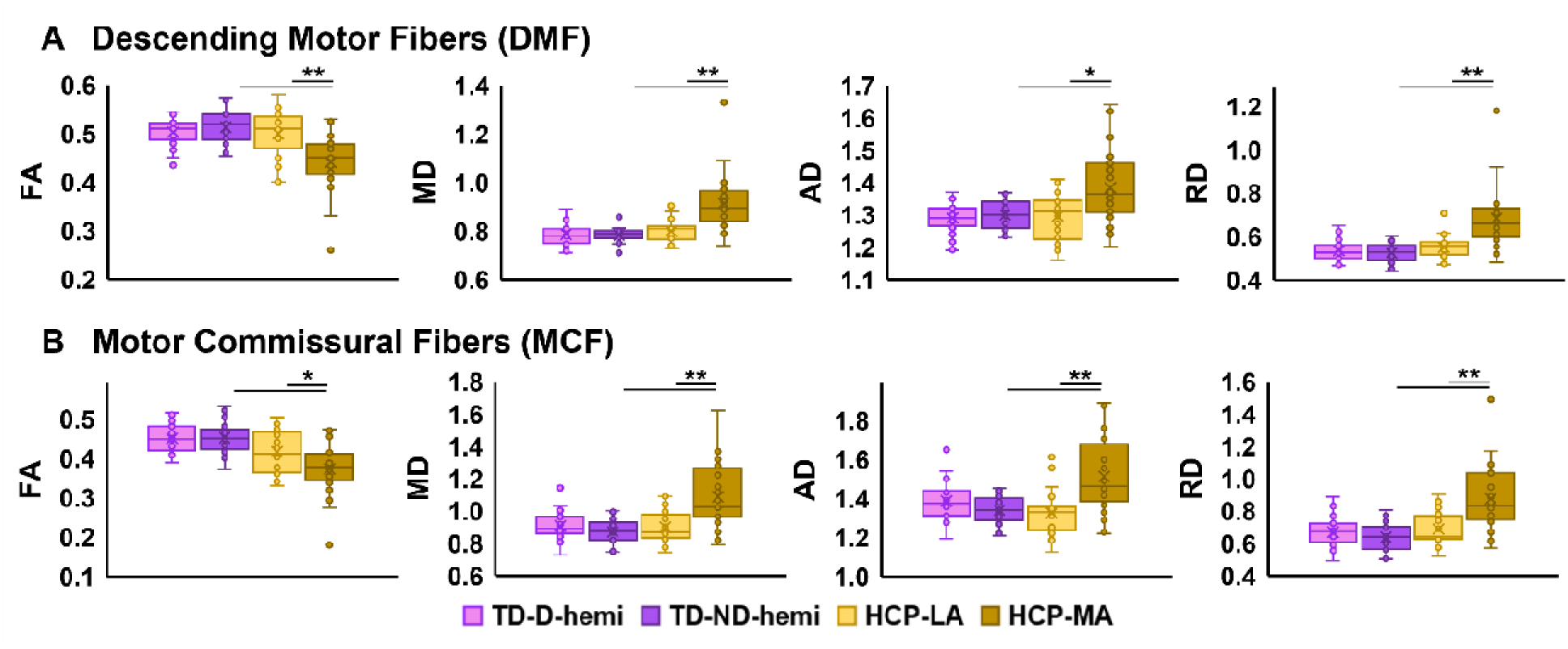
Diffusion MRI measures of motor-pathway microstructure. Box-and-whisker plots show fractional anisotropy (FA), mean diffusivity (MD), axial diffusivity (AD) and radial diffusivity (RD) in **(A)** descending motor fibers (DMF) and **(B)** motor commissural fibers (MCF). Data are shown for th dominant (D-hemi) and nondominant (ND-hemi) hemispheres of typically developing children (TD; n = 23) and the less affected (LA; n = 25) and more affected (MA; n = 23) hemispheres of children with hemiplegic cerebral palsy (HCP). Tracts were reconstructed using functionally defined primary motor cortex regions combined with anatomical brainstem and corpus callosum regions for DMF and MCF, respectively. Boxes represent the interquartile range, horizontal lines indicate medians, crosses denote means and circles represent individual observations. Within-participant comparisons used Wilcoxon signed-rank tests, and between-group comparisons used Mann–Whitney U tests with false-discovery-rate correction. Horizontal brackets indicate significant comparisons: ∗ < 0.05, ∗∗ < 0.01.

MCF showed a similar pattern (Fig. 6B): the MA hemisphere had lower FA (*P* < 0.01, r > 0.67) and higher MD (*P* < 0.001, r > 0.76), AD (*P* < 0.001, r > 0.60), and RD (*P* < 0.001, r > 0.77) than the TD group and the LA hemisphere of the HCP group. MCF integrity did not differ between CSC and PV groups. However, within the PV group, MA vs. LA differences were observed for the MD, AD, and RD of the MCF (*P* < 0.05, r > 0.79; Supplementary Fig. 3).

### Intermodality associations

Larger lesion/ventricular dilation volume was associated with higher rMT (Rho = 0.77, *P* < 0.05) and poorer range of motion, accuracy, and dexterity (–0.74 < Rho < –0.57, *P* < 0.05) (Table 2). Across groups, hand, and hemispheres, better dexterity was associated with higher FA (Rho = 0.52, *P* < 0.001) and lower MD and RD (–0.51 < Rho < –0.48, *P* < 0.001) of the DMFs. ERD_mu_ was correlated with MD and RD (0.58 < Rho < 0.62, *P* < 0.05), and ERD_beta_ correlated with the DMF AD (Rho = 0.55, *P* < 0.05). Lower LI_TMS_ was associated with poorer MCF integrity including lower FA (Rho = 0.41, *P* < 0.05) and MD and RD (–0.45 < Rho < –0.43, *P* < 0.01).

**Table 2.** Inter-modality correlations among structural, functional, and behavioral metrics.

| **Metric #1** | **Metric #2** | **Spearman's *Rho*** | **P-value (FDR)** | **Interpretation** |
|---|---|---|---|---|
| Lesion size (HCP only) | rMT | 0.77 | 0.021 | Structure - physiology |
| Lesion size (HCP only) | ROM | –0.58 | 0.029 | Structure - behavior |
| Lesion size (HCP only) | Accuracy | –0.57 | 0.030 | Structure - behavior |
| Lesion size (HCP only) | Dexterity | –0.74 | 0.001 | Structure - behavior |
| DMF FA | Dexterity | 0.52 | 0.0001 | Structure - behavior |
| DMF MD/RD | Dexterity | –0.48 to –51 | < 0.0001 | Structure - behavior |
| DMF FA | Reaction time | –0.46 | 0.0001 | Structure - behavior |
| DMF MD/RD | Reaction time | 0.50 to 0.51 | < 0.0001 | Structure - behavior |
| DMF MD/RD | ERD<sub>mu</sub> | 0.52 to 0.63 | < 0.05 | Structure - physiology |
| DMF AD | ERD<sub>beta</sub> | 0.55 | 0.025 | Structure - physiology |
| MCF FA | LI<sub>TMS</sub> | 0.43 | 0.0005 | Structure - physiology |
| MCF MD/RD | LI<sub>TMS</sub> | –0.43 to –0.48 | < 0.001 | Structure - physiology |
| ERD<sub>beta</sub> | Accuracy | –0.67 | 0.045 | Physiology - behavior |
| LI<sub>TMS</sub> | LI<sub>MRR</sub> | 0.44 | 0.048 | Physiology - physiology |
| LI<sub>TMS</sub> | ROM | 0.65 | 0.0001 | Physiology - behavior |
| LI<sub>TMS</sub> | Accuracy | 0.45 | 0.002 | Physiology - behavior |
| LI<sub>TMS</sub> | Dexterity | 0.65 | 0.0001 | Physiology - behavior |
| LI<sub>TMS</sub> | Fluency | 0.70 | 0.0001 | Physiology - behavior |

AD = axial diffusivity; ERD = event-related desynchronization; FA = fractional anisotropy; HCP = hemiplegic cerebral palsy; LI = lateralization index; MD = mean diffusivity; MRR = movement-related response; PH = paretic hand; RD = radial diffusivity; rMT = resting motor threshold; ROM = range of motion; TMS = transcranial magnetic stimulation.
P-values obtained from Spearman's correlation and corrected for multiple comparisons (FDR).

Within the HCP group, lower LI_TMS_ following the LA hemisphere stimulation was correlated with lower LI_MRR_ (Rho = 0.44, *P* < 0.05), indicating concordant alterations in the corticospinal organization and voluntary cortical recruitment. Lower LI_TMS_ was also associated with poorer range of motion, accuracy, dexterity, and fluency (0.43 < Rho < 0.61, *P* < 0.05). Reduced contralateral ERD_beta_ was associated with poorer P-hand movement accuracy (Rho = – 0.67, *P* < 0.05).

A cross-validated partial least-squares structural-functional integrity score significantly predicted motor function (Q² = 0.42, *P* ≤ 0.001; Supplementary Table 1). DMF and MCF integrity also predicted motor performance (Q² = 0.25 and 0.19, *P* < 0.01) and were moderately correlated (r = 0.63, *P* < 0.01). The full structural-functional model outperformed the structural model.

Finally, ERD_beta_ mediated the association between the DMF microstructural properties and overall motor function (indirect effect for FA: 0.12 SD, 95% CI [0.03, 0.24], MD: –0.11 [– 0.23, –0.03], AD: –0.11 [–0.25, 0.02], RD: –0.11 [–0.21, –0.02]).

## Discussion

Using multimodal neuroimaging, we show that developmental motor neuroplasticity in children with HCP is a coordinated structure–physiology–behavior process rather than a direct consequence of structural injury alone. Early unilateral injury disrupts descending and commissural motor pathways, altering corticospinal organization, redistributing cortical motor recruitment, and impairing upper-extremity function. This reorganization is not confined to the MA hemisphere: each hemisphere shows distinct alterations in movement-related oscillatory dynamics, indicating that early injury reshapes the bilateral motor network. Mediation analysis further ordered these levels hierarchically, with structural remodeling constraining corticospinal physiology and thereby shaping cortical recruitment and motor behavior. These findings provide a mechanistic framework for understanding individual variability in motor outcomes and support rehabilitation and neuromodulation strategies accounting for structural constraints and patient-specific bilateral motor-network reorganization.

### Structural remodeling establishes the substrate for developmental motor plasticity

Structural remodeling in HCP was evident at macroscopic and microstructural levels. Macroscopically, lesion type was associated with distinct patterns of motor system organization.^8,11^ All children with PV lesions retained an anatomically preserved M1,^8^ with movement-related source localization closely matching the Brodmann area 4 parcellation, despite DMFs and MCFs displacement around the enlarged ventricle. In contrast, CSC lesions directly involved the sensorimotor cortex in a subset of children, displacing functionally localized M1 and reorganizing descending motor pathways. Therefore, injury preceding corticospinal maturation appears to spare the representation while deforming the pathways around it, whereas later injury involving cortex relocates the representation itself and requires reconstruction of descending and interhemispheric tracts which may fail in some children with larger CSC lesions.^8,11^

Seeding tractography from functionally localized M1 rather than atlas-defined landmarks identified lesion-specific patterns. Conventional tractography reconstructs the DMFs from anatomically defined cortical and subcortical regions of interest and therefore assumes preserved cortical organization,^20,21^ an assumption that may not hold in HCP, where motor representations can be displaced after early injury.^5,8,54^ TMS-defined M1 offers an alternative cortical seed, but reliable motor mapping is often difficult in children with extensive cortical lesions, because direct M1 involvement and elevated rMT can preclude a reproducible motor hotspot.^55^ Seeding from M1 localized with electromagnetic source imaging instead allowed us to reconstruct the descending and commissural pathways engaged during voluntary upper-extremity movement. This strategy characterizes physiologically relevant pathway reorganization.^5,29^

Children with HCP also exhibited microstructural abnormalities in MA DMF and MCF, indicating effects on corticospinal output and interhemispheric connectivity. Reduced FA and increased MD, AD, and RD are compatible with disrupted fiber organization and tissue integrity, potentially reflecting axonal loss, gliosis, cystic degeneration, altered myelination, or cerebral edema.^21,56–55^ Rather than attributing individual diffusion metrics to specific histopathology, we interpret them as convergent evidence of altered microstructural organization. These findings extend prior DWI studies^21,24,25^ localizing such abnormalities to pathways that support voluntary movement.

DMF abnormalities were present in the MA hemisphere of both lesion groups but were most pronounced after CSC injury, consistent with damage occurring at a more advanced corticospinal development stage.^5,22^ MCF metrics showed no overall lesion-type difference, although children with PV lesions demonstrated hemispheric asymmetry in diffusivity measures. Timing of injury may therefore influence descending and commissural pathways differently: earlier injury may permit asymmetric transcallosal connections development, whereas later cortical injury may compromise both corticospinal and transcallosal architecture more severely.

### Cortical recruitment reflects bilateral motor-system reorganization

Movement-related cortical activity showed that developmental plasticity extends beyond the injured hemisphere. During P-hand movement, children with HCP showed reduced movement-related lateralization (Fig. 3C), consistent with bilateral network recruitment. Unlike post-stroke bilateral activation in adults, this organization likely emerges during corticospinal maturation and becomes integrated into the developing motor system.^57^

Abnormalities were also evident during NP-hand movement. Despite relatively preserved motor performance, children with HCP showed slower responses (Fig. 3A), reduced theta synchronization (Fig. 4A) and greater beta lateralization (Supplementary Fig. 2). The LA hemisphere is therefore not functionally typical; it appears to participate in a reorganized bilateral system supporting motor planning and execution of both upper extremities.^6^

Oscillatory abnormalities further revealed frequency- and hemisphere-specific alterations in motor-system dynamics. Movement-related ERD_mu_ and ERD_beta_ are well-established electrophysiological signatures of sensorimotor cortical engagement during movement preparation and execution, reflecting ongoing rhythmic activity suppression within recruited cortical networks.^28,53^ In children with cerebral palsy, alterations in these oscillations have been linked to atypical motor planning, movement execution, and lower-extremity performance.^34,58^

Here, children with HCP showed attenuated ERD_mu_ and ERD_beta_ in the ipsilesional hemisphere during P-hand movement (Fig. 4B), and these alterations tracked reduced DMF integrity (Table 2). Thus, descending motor-pathway disruption accompanies altered modulation of ipsilesional sensorimotor cortical activity during voluntary movement. Reduced contralateral ERD_beta_ was associated with poorer movement accuracy.

In contrast, ERS_theta_ was selectively reduced during NP-hand movement in the contralesional hemisphere (Fig. 4A) and was unrelated to DMF or MCF integrity and to motor performance. Unlike mu and beta rhythms, which track movement-related sensorimotor activation, theta activity is implicated in motor planning and in integrating sensory information, such as visual feedback, with intended actions.^59,60^ Its selective alteration may therefore reflect a component of bilateral motor-network reorganization that is not directly determined by corticospinal structural integrity. Developmental motor reorganization after early unilateral brain injury thus involves frequency-specific changes across the bilateral motor network, spanning movement preparation, sensorimotor engagement, and execution.^61^

Increased beta lateralization during NP-hand movement suggests greater reliance on the LA hemisphere for descending motor output, whereas reduced gamma lateralization during P-hand movement indicates less contralaterally restricted execution-related processing (Supplementary Fig. 2).^62,63^ Gamma peak latency and frequency showed similar trends without reaching significance, likely reflecting substantial interindividual variability.^64^

Slower response times for both hands suggest that bilateral reorganization affects movement preparation and initiation.^28^ Higher inhibition accuracy during P-hand trials (Fig. 3B) may reflect slower motor preparation rather than superior inhibitory control, consistent with a speed-accuracy trade-off during developmental motor reorganization.

### Corticospinal physiology links structural integrity to motor function

nTMS revealed physiological reorganization of corticospinal motor output after early unilateral injury. Most children with HCP retained ipsilateral corticospinal projections from the LA hemisphere (Fig. 5C), consistent with altered developmental pruning.^7,8^ The MA hemisphere showed higher rMT (Fig. 5B), indicating reduced corticospinal excitability, while ipsilateral MEPs showed prolonged latencies (Fig. 5A), suggesting less efficient conduction through reorganized descending pathways.

Corticospinal physiology tracked both lesion burden and motor function (Table 2). Larger lesions were associated with higher rMT, and reduced corticospinal lateralization correlated with poorer performance across all movement-quality domains. Corticospinal lateralization also correlated with MCF integrity, indicating that efficient descending motor output depends on both corticospinal preservation and interhemispheric connectivity. Commissural-pathway disruption therefore may contribute to interhemispheric physiological imbalance and impaired coordinated motor output.

Although movement-related cortical responses and nTMS both showed reduced hemispheric lateralization, their association was only moderate, likely because nTMS assesses corticospinal output capacity whereas movement-related responses reflect distributed cortical network recruitment during voluntary movement.^65,66^ Corticospinal organization therefore constrains, but does not fully determine, voluntary cortical recruitment, leaving higher-order motor networks partially free to adapt during development.

### Structural and microstructural alterations constrain functional outcome

Relationships between structural measures and motor performance reveal constraints imposed by early brain injury. Lesion extent was associated with upper-extremity motor function (Table 2), indicating that macroscopic structural disruption influences motor capacity. DMF integrity was also associated with dexterity, indicating that preserved descending pathways provide an anatomical substrate for upper-extremity control. Motor outcome is therefore shaped by both lesion extent and descending pathway integrity, reinforcing corticospinal integrity as a major upper-extremity function determinant.^13,22,67^ Preserved white-matter architecture within the LA hemisphere accompanied extensive physiological reorganization, indicating that structural integrity provides the anatomical framework for motor control and partially shapes functional organization.^68,69^ Together, these findings suggest that the developing nervous system adapts its function within the injury-imposed structural constraints.

### An integrated structure–physiology–behavior cascade explains motor dysfunction

Although each modality identified motor-system abnormalities, this study demonstrates how these findings converge into a unified biological framework. Structural connectivity forms the anatomical level of this hierarchy, corticospinal physiology and cortical recruitment the successive functional levels, and motor behavior the resulting functional outcome. These levels appear interconnected within developmental motor reorganization rather than independent consequences of early brain injury. The correlation analyses, latent structural-functional model, and mediation analysis provide convergent evidence supporting this hierarchical organization.

Structural injury established the anatomical framework of the cascade. Larger lesion size and diminished DMF integrity were associated with poorer movement quality, and DMF microstructure correlated with ERD_mu_ and ERD_beta_ (Table 2). DMF integrity may therefore influence both performance and sensorimotor-cortical recruitment during voluntary movement.^70,71^ MCF integrity, in contrast, was more closely related to corticospinal lateralization (Table 2), supporting its role in interhemispheric physiological balance.^21,72,73^

Corticospinal organizations link structural remodeling to cortical function. The association between movement-related and TMS-derived lateralization suggests that corticospinal organization constrains voluntary cortical recruitment while permitting higher-order cortical adaptation,^69^ which may help explain why children with similar lesions show different cortical recruitment strategies and motor outcomes.^8,68^

Multivariate analyses reinforced this model. DMF and MCF integrity independently predicted motor performance, yet the combined structural-functional model predicted movement quality better than structural measures alone. Beta-band modulation also mediated the association between DMF integrity and overall motor performance, suggesting that white-matter disruption affects behavior partly through altered cortical recruitment during movement. Causal inference cannot be established in this cross-sectional design, but the results support this relationship as a biologically plausible mechanism.

Collectively, these findings support a model in which early unilateral injury disrupts descending and commissural pathways, prompting reorganization of corticospinal output and cortical motor recruitment,^8,32,34^ that ultimately shapes upper-extremity performance bilaterally.^5,16^ Structural connectivity, corticospinal physiology, cortical activity, and behavior should therefore be viewed as complementary measures of a single developmental adaptive process.

### Clinical implications

Our findings highlight a major gap between neuroimaging evidence on developmental plasticity and clinical care for children with HCP. Clinicians cannot reliably predict individual long-term rehabilitation approach. Lesion characteristics and corticospinal tract integrity are informative but capture only part of the motor function biology. Our results indicate that structural connectivity, corticospinal physiology, and cortical dynamics provide complementary information about motor performance: children with similar lesion profiles may differ in outcome because they differ in physiological adaptation and motor-network reorganization. Multimodal assessment could identify individual neuroplastic profiles to inform prognostic counseling, rehabilitation selection, and treatment timing.

Our findings also have direct implications for development of therapeutic neuromodulation for people with early unilateral brain injury. Suppression of the LA hemisphere commonly assumes that contralesional recruitment is maladaptive. Yet, the LA hemisphere showed substantial physiological reorganization and often generated corticospinal output to both upper extremities (Fig. 5C). Although greater ipsilateral corticospinal contribution was associated with poorer upper-extremity motor performance, it may suggest that contralesional corticospinal recruitment may contribute to motor function rather than simply compete with the MA hemisphere. Interventions targeting interhemispheric balance should therefore be guided by each child’s motor-network organization rather than lesion characteristics alone, and account for altered hemodynamic, metabolic, and electrophysiological activity.^74–78^ Integrating diffusion MRI, nTMS, and functional neurophysiology could support patient stratification and individualized rehabilitation neuromodulation.

### Limitations and future directions

This cross-sectional design cannot establish the developmental trajectory of, or causal relationships among, structural connectivity, corticospinal physiology, cortical recruitment, and motor function. Longitudinal studies are needed to determine how these biomarkers evolve with development and respond to rehabilitation. We did not directly assess interhemispheric inhibition or effective connectivity, which may influence motor reorganization; combining diffusion MRI with paired-pulse TMS, TMS–EEG, and network-connectivity analyses could better define how structural remodeling shapes bilateral motor-network communication. Some physiological findings require replication. Theta synchronization was largely unrelated to structural injury and motor performance (Fig. 4), suggesting that it may reflect higher-order motor planning not captured by conventional clinical assessments. Gamma oscillations showed altered lateralization with non-significant trends in peak latency and frequency (Supplementary Fig. 2), but substantial inter-individual variability limited statistical power. Adding quantitative mirror-movement measures, bimanual coordination, and motor learning may clarify the functional significance of these oscillatory biomarkers.

## Conclusion

Developmental motor neuroplasticity in HCP is best understood as a coordinated structure– physiology–behavior process. Early unilateral injury disrupts descending and commissural motor pathways, constraining corticospinal organization and cortical recruitment during voluntary movement. The developing brain then redistributes motor processing across a bilateral network, partially preserving upper extremity motor function. By integrating diffusion MRI, movement-related cortical activity, oscillatory dynamics, nTMS, and standardized motor assessments in the same cohort, this study identifies complementary multimodal candidate biomarkers that capture distinct motor-system reorganization levels and link structural injury to motor function. This multimodal approach moves beyond lesion burden alone by providing an individualized profile of structural connectivity, corticospinal organization, and cortical recruitment. Such profiles could improve prognostication, identify neuroplastic patterns relevant to treatment selection, and guide more targeted rehabilitation and neuromodulation strategies in children with HCP.

## Acknowledgement

We thank Dr. Sabrina Shandley and Eryn Armstrong for their contribution in behavioral assessment.

## Data availability

The data are available from the corresponding author upon request.

## Supplementary material

Supplementary material is available online.

## References

1. Rosenbaum P, Paneth N, Leviton A, et al. A report: the definition and classification of cerebral palsy April 2006. Dev Med Child Neurol Suppl. 2007;109(suppl 109):8–14.

2. Gordon AM, Bleyenheuft Y, Steenbergen B. Pathophysiology of impaired hand function in children with unilateral cerebral palsy. Dev Med Child Neurol. 2013;55 Suppl 4:32–37. doi:10.1111/dmcn.12304

3. Arner M, Eliasson AC, Nicklasson S, Sommerstein K, Haggland G. Hand function in cerebral palsy. Report of 367 children in a population-based longitudinal health care program. J Hand Surg [Am]. 2008;33(8):1337–1347. doi:10.1016/j.jhsa.2008.02.032

4. Wiklund LM, Uvebrant P. Hemiplegic cerebral palsy: correlation between CT morphology and clinical findings. Dev Med Child Neurol. 1991;33(6):512–523. doi:10.1111/j.1469-8749.1991.tb14916.x

5. Shahdadian S, Song Y, Vaysi A, et al. Functional and structural neuroplasticity of somatosensory system in hemiplegic cerebral palsy. Brain Commun. 2026;8(3):fcag185.

6. Reid LB, Rose SE, Boyd RN. Rehabilitation and neuroplasticity in children with unilateral cerebral palsy. Nat Rev Neurol. 2015;11(7):390–400. doi:10.1038/nrneurol.2015.97

7. Staudt M. (Re-)organization of the developing human brain following periventricular white matter lesions. Neurosci Biobehav Rev. 2007;31(8):1150–1156. doi:10.1016/j.neubiorev.2007.05.005

8. Staudt M. Reorganization after pre- and perinatal brain lesions. J Anat. 2010;217(4):469–474. doi:10.1111/j.1469-7580.2010.01262.x

9. Fehlings D, Krishnan P, Ragguett RM, et al. Neurodevelopmental profiles of children with unilateral cerebral palsy associated with middle cerebral artery and periventricular venous infarctions. Dev Med Child Neurol. 2021;63(6):729–735.

10. Kolb B, Mychasiuk R, Muhammad A, Gibb R. Chapter 2 - Brain Plasticity in the Developing Brain. In: Merzenich MM, Nahum M, Vleet TMV, eds. Changing Brains. Vol 207. Progress in Brain Research. Elsevier; 2013:35–64. 10.1016/B978-0-444-63327-9.00005-9

11. Fiori S, Guzzetta A. Plasticity following early-life brain injury: insights from quantitative MRI. In: Seminars in Perinatology. Vol 39. Elsevier; 2015:141–146.

12. Eyre JA. Corticospinal tract development and its plasticity after perinatal injury. Neurosci Biobehav Rev. 2007;31(8):1136–1149.

13. Holmström L, Vollmer B, Tedroff K, et al. Hand function in relation to brain lesions and corticomotor-projection pattern in children with unilateral cerebral palsy. Dev Med Child Neurol. 2010;52(2):145–152.

14. Kuhtz-Buschbeck JP, Sundholm LK, Eliasson AC, Forssberg H. Quantitative assessment of mirror movements in children and adolescents with hemiplegic cerebral palsy. Dev Med Child Neurol. 2000;42(11):728–736.

15. Adler C, Berweck S, Lidzba K, Becher T, Staudt M. Mirror movements in unilateral spastic cerebral palsy: Specific negative impact on bimanual activities of daily living. Eur J Paediatr Neurol. 2015;19(5):504–509.

16. Feys H, Eyssen M, Jaspers E, et al. Relation between neuroradiological findings and upper limb function in hemiplegic cerebral palsy. Eur J Paediatr Neurol. 2010;14(2):169–177. doi:10.1016/j.ejpn.2009.01.004

17. Jiang H, Li X, Jin C, et al. Early diagnosis of spastic cerebral palsy in infants with periventricular white matter injury using diffusion tensor imaging. Am J Neuroradiol. 2019;40(1):162–168.

18. Staudt M, Gerloff C, Grodd W, Holthausen H, Niemann G, Krägeloh-Mann I. Reorganization in congenital hemiparesis acquired at different gestational ages. Ann Neurol Off J Am Neurol Assoc Child Neurol Soc. 2004;56(6):854–863.

19. Simon-Martinez C, Jaspers E, Mailleux L, et al. Corticospinal tract wiring and brain lesion characteristics in unilateral cerebral palsy: determinants of upper limb motor and sensory function. Neural Plast. 2018;2018(1):2671613.

20. Jaspers E, Byblow WD, Feys H, Wenderoth N. The Corticospinal Tract: A Biomarker to Categorize Upper Limb Functional Potential in Unilateral Cerebral Palsy. Front Pediatr. 2016;Volume 3-2015. doi:10.3389/fped.2015.00112

21. Papadelis C, Ahtam B, Feldman HA, et al. Altered White Matter Connectivity Associated with Intergyral Brain Disorganization in Hemiplegic Cerebral Palsy. Neuroscience. 2019;399:146–160. doi:10.1016/j.neuroscience.2018.12.028

22. Papadelis C, Kaye H, Shore B, Snyder B, Grant PE, Rotenberg A. Maturation of corticospinal tracts in children with hemiplegic cerebral palsy assessed by diffusion tensor imaging and transcranial magnetic stimulation. Front Hum Neurosci. 2019;13:254.

23. Jaatela J, Nurmi T, Vallinoja J, Maenpaa H, Sairanen V, Piitulainen H. Altered corpus callosum structure in adolescents with cerebral palsy: Connection to gait and balance. Brain Struct Funct. 2023;228(8):1901–1915.

24. Gupta D, Barachant A, Gordon AM, et al. Effect of sensory and motor connectivity on hand function in pediatric hemiplegia. Ann Neurol. 2017;82(5):766–780.

25. Jaatela J, Nurmi T, Vallinoja J, Mäenpää H, Sairanen V, Piitulainen H. Altered corpus callosum structure in adolescents with cerebral palsy: connection to gait and balance. Brain Struct Funct. 2023;228(8):1901–1915.

26. Robert MT, Gutterman J, Ferre CL, et al. Corpus callosum integrity relates to improvement of upper-extremity function following intensive rehabilitation in children with unilateral spastic cerebral palsy. Neurorehabil Neural Repair. 2021;35(6):534–544.

27. Pfurtscheller G, Silva FHL da. Event-related EEG/MEG synchronization and desynchronization: basic principles. Clin Neurophysiol. 1999;110(11):1842–1857. 10.1016/S1388-2457(99)00141-8

28. Cheyne DO. MEG studies of sensorimotor rhythms: A review. Exp Neurol. 2013;245:27–39. 10.1016/j.expneurol.2012.08.030

29. Papadelis C, Butler EE, Rubenstein M, et al. Reorganization of the somatosensory cortex in hemiplegic cerebral palsy associated with impaired sensory tracts. NeuroImage Clin. 2018;17:198–212.

30. Pihko E, Nevalainen P, Vaalto S, et al. Reactivity of sensorimotor oscillations is altered in children with hemiplegic cerebral palsy: A magnetoencephalographic study. Hum Brain Mapp. 2014;35(8):4105–4117. doi:10.1002/hbm.22462

31. Kurz MJ, Wilson TW, Arpin DJ, others. Motor beta cortical oscillations are related with the gait kinematics in children with cerebral palsy. Eur J Neurosci. 2020;52(10):4123–4135.

32. Hinton EH, Wilson TW, Kurz MJ. Adults with cerebral palsy exhibit uncharacteristic cortical oscillations during an adaptive sensorimotor control task. J Neurophysiol. 2024;131(5):1234–1245.

33. Vallinoja J, Maenpaa H, Sairanen V, others. A multimodal study using fMRI and MEG in cerebral palsy. Brain Connect. 2024;14(2):100–115.

34. Kurz MJ, Becker KM, Heinrichs-Graham E, Wilson TW. Neurophysiological abnormalities in the sensorimotor cortices during the motor planning and movement execution stages of children with cerebral palsy. Dev Med Child Neurol. 2014;56(11):1072–1077.

35. Rossini PM, Burke D, Chen R, et al. Non-invasive electrical and magnetic stimulation of the brain, spinal cord, roots and peripheral nerves: Basic principles and procedures for routine clinical and research application. An updated report from an IFCN Committee. Clin Neurophysiol. 2015;126(6):1071–1107.

36. Groppa S, Oliviero A, Eisen A, et al. A practical guide to diagnostic transcranial magnetic stimulation: report of an IFCN committee. Clin Neurophysiol. 2012;123(5):858–882.

37. Sondergaard RE, Martino D, Kiss ZH, Condliffe EG. TMS motor mapping methodology and reliability: a structured review. Front Neurosci. 2021;15:709368.

38. Krieg SM, Lioumis P, Mäkelä JP, et al. Protocol for motor and language mapping by navigated TMS in patients and healthy volunteers; workshop report. Acta Neurochir (Wien). 2017;159(7):1187–1195.

39. Feys H, De Weerdt W, Verheyden G, others. Functional recovery in hemiplegic cerebral palsy: Ipsilateral electromyographic responses to focal transcranial magnetic stimulation. Dev Med Child Neurol. 1998;40(4):236–242.

40. Maegaki Y, Yamamoto T, Yamashita Y, others. Functional reorganization of motor areas in hemiplegic cerebral palsy: A PET study. Dev Med Child Neurol. 1997;39(7):431–439.

41. Eyre JA, Taylor JP, Villagra F, Smith M, Miller S. Evidence of activity-dependent withdrawal of corticospinal projections during human development. Neurology. 2001;57(9):1543–1554. doi:10.1212/WNL.57.9.1543

42. Randall M, Johnson L, Reddihough D. The melbourne assessment 2. Melb R Child Hosp. Published online 1999.

43. Corona Ludovica PC Rijal Sakar, Tanritanir Omer, Shahdadian Sadra, Keator Cynthia G, Tran Linh, Malik Saleem I, Bosemani Madhan, Hansen Daniel, Shahani Dave, Perry M Scott. Electromagnetic Source Imaging in Presurgical Evaluation of Children with Drug-Resistant Epilepsy. J Vis Exp. 2024;(211):e66494. doi:doi:10.3791/66494

44. Tadel F, Baillet S, Mosher JC, Pantazis D, Leahy RM. Brainstorm: A user-friendly application for MEG/EEG analysis. Comput Intell Neurosci. 2011;2011(1):879716.

45. Dale AM, Liu AK, Fischl BR, et al. Dynamic Statistical Parametric Mapping: Combining fMRI and MEG for High-Resolution Imaging of Cortical Activity. Neuron. 2000;26(1):55–67. doi:10.1016/S0896-6273(00)81138-1

46. Hämäläinen MS, Ilmoniemi RJ. Interpreting magnetic fields of the brain: minimum norm estimates. Med Biol Eng Comput. 1994;32:35–42.

47. Gramfort A, Papadopoulo T, Olivi E, Clerc M. OpenMEEG: opensource software for quasistatic bioelectromagnetics. Biomed Eng Online. 2010;9:1–20.

48. Cheyne D, Bakhtazad L, Gaetz W. Spatiotemporal mapping of cortical activity accompanying voluntary movements using an event-related beamforming approach. Hum Brain Mapp. 2006;27(3):213–229.

49. Pfurtscheller G. Event-related synchronization (ERS): an electrophysiological correlate of cortical areas at rest. Electroencephalogr Clin Neurophysiol. 1992;83(1):62–69.

50. Seghier ML. Laterality index in functional MRI: methodological issues. Magn Reson Imaging. 2008;26(5):594–601.

51. Yeh FC, Wedeen VJ, Tseng WYI. Generalized ${q} $-sampling imaging. IEEE Trans Med Imaging. 2010;29(9):1626–1635.

52. Yeh FC, Panesar S, Barrios J, et al. Automatic removal of false connections in diffusion MRI tractography using topology-informed pruning (TIP). Neurotherapeutics. 2019;16(1):52–58.

53. Neuper C, Pfurtscheller G. Event-related dynamics of cortical rhythms: frequency-specific features and functional correlates. Int J Psychophysiol. 2001;43(1):41–58.

54. Thomas C, Ye FQ, Irfanoglu MO, et al. Anatomical accuracy of brain connections derived from diffusion MRI tractography is inherently limited. Proc Natl Acad Sci. 2014;111(46):16574–16579.

55. Groppa S, Oliviero A, Eisen A, et al. A practical guide to diagnostic transcranial magnetic stimulation: report of an IFCN committee. Clin Neurophysiol. 2012;123(5):858–882.

56. Pierpaoli C, Jezzard P, Basser PJ, Barnett A, Di Chiro G. Diffusion tensor MR imaging of the human brain. Radiology. 1996;201(3):637–648.

57. Cauraugh JH, Summers JJ. Neural plasticity and bilateral movements: A rehabilitation approach for chronic stroke. Prog Neurobiol. 2005;75(5):309–320. 10.1016/j.pneurobio.2005.04.001

58. Kurz MJ, Proskovec AL, Gehringer JE, Heinrichs-Graham E, Wilson TW. Children with cerebral palsy have altered oscillatory activity in the motor and visual cortices during a knee motor task. NeuroImage Clin. 2017;15:298–305.

59. Tomassini A, Ambrogioni L, Medendorp WP, Maris E. Theta oscillations locked to intended actions rhythmically modulate perception. Elife. 2017;6:e25618.

60. Dufour B, Thénault F, Bernier PM. Theta-band EEG activity over sensorimotor regions is modulated by expected visual reafferent feedback during reach planning. Neuroscience. 2018;385:47–58.

61. Papadelis C, Arfeller C, Erla S, Nollo G, Cattaneo L, Braun C. Inferior frontal gyrus links visual and motor cortices during a visuomotor precision grip force task. Brain Res. 2016;1650:252–266.

62. Nowak M, Zich C, Stagg CJ. Motor cortical gamma oscillations: what have we learnt and where are we headed? Curr Behav Neurosci Rep. 2018;5(2):136–142.

63. Cheyne D, Bells S, Ferrari P, Gaetz W, Bostan AC. Self-paced movements induce high-frequency gamma oscillations in primary motor cortex. NeuroImage. 2008;42(1):332–342. doi:10.1016/j.neuroimage.2008.04.178

64. Cheyne D, Ferrari P. MEG studies of motor cortex gamma oscillations: Evidence for a gamma “fingerprint” in the brain? Front Hum Neurosci. 2013;(SEP). doi:10.3389/fnhum.2013.00575

65. Krings T, Buchbinder BR, Butler WE, et al. Functional magnetic resonance imaging and transcranial magnetic stimulation: complementary approaches in the evaluation of cortical motor function. Neurology. 1997;48(5):1406–1416.

66. Staudt M, Grodd W, Gerloff C, Erb M, Stitz J, Krägeloh-Mann I. Two types of ipsilateral reorganization in congenital hemiparesis: a TMS and fMRI study. Brain J Neurol. 2002;125(Pt 10):2222–2237. doi:10.1093/brain/awf227

67. Mailleux L, Franki I, Emsell L, et al. The relationship between neuroimaging and motor outcome in children with cerebral palsy: A systematic review—Part B diffusion imaging and tractography. Res Dev Disabil. 2020;97:103569.

68. Simon-Martinez C, Decraene L, Zielinski I, et al. The impact of brain lesion characteristics and the corticospinal tract wiring on mirror movements in unilateral cerebral palsy. Sci Rep. 2022;12(1):16301.

69. Simon-Martinez C, Jaspers E, Alaerts K, et al. Influence of the corticospinal tract wiring pattern on sensorimotor functional connectivity and clinical correlates of upper limb function in unilateral cerebral palsy. Sci Rep. 2019;9(1):8230.

70. Kirton A, DeVeber G, Pontigon AM, Macgregor D, Shroff M. Presumed perinatal ischemic stroke: vascular classification predicts outcomes. Ann Neurol Off J Am Neurol Assoc Child Neurol Soc. 2008;63(4):436–443.

71. De Vries LS, Groenendaal F, Van Haastert IC, Eken P, Rademaker KJ, Meiners LC. Asymmetrical myelination of the posterior limb of the internal capsule in infants with periventricular haemorrhagic infarction: an early predictor of hemiplegia. Neuropediatrics. 1999;30(06):314–319.

72. Weinstein M, Green D, Geva R, et al. Interhemispheric and intrahemispheric connectivity and manual skills in children with unilateral cerebral palsy. Brain Struct Funct. 2014;219(3):1025–1040.

73. Mailleux L, Simon-Martinez C, Radwan A, et al. White matter characteristics of motor, sensory and interhemispheric tracts underlying impaired upper limb function in children with unilateral cerebral palsy. Brain Struct Funct. 2020;225(5):1495–1509.

74. Damirchi BG, Jahromi S, Vaysi A, Partamian H, Shahdadian S, Papadelis C. Multimodal Validation of Temporal Interference in a 3D-Printed Pediatric Head Phantom. NeuroImage. Published online 2026:122141.

75. Damirchi BG, Jahromi S, Sdoukopoulou G, Shahdadian S, Papadelis C. Induction of gamma oscillations via transcranial temporal interference stimulation: a human head phantom study. In: 2025 47th Annual International Conference of the IEEE Engineering in Medicine and Biology Society (EMBC). IEEE; 2025:1–4.

76. Shahdadian S, Wang X, Liu H. Directed physiological networks in the human prefrontal cortex at rest and post transcranial photobiomodulation. Sci Rep. 2024;14(1):10242.

77. Shahdadian S, Wang X, Wanniarachchi H, Chaudhari A, Truong NCD, Liu H. Neuromodulation of brain power topography and network topology by prefrontal transcranial photobiomodulation. J Neural Eng. 2022;19(6):066013.

78. Wang X, Ma LC, Shahdadian S, Wu A, Truong NCD, Liu H. Metabolic connectivity and hemodynamic-metabolic coherence of human prefrontal cortex at rest and post photobiomodulation assessed by dual-channel broadband NIRS. Metabolites. 2022;12(1):42.

