## Supplementary materials for "Functional and structural neuroplasticity of motor system in hemiplegic cerebral palsy"

Assistant Vice President of Neuroscience Research, Cook Children's Health Care System

1500 Cooper St., Fort Worth, TX, 76104, USA

ORCID: 0000-0001-6125-9217


**Short title**: Mapping Motor Plasticity in Hemiplegia


**Keywords:** cortical oscillations, corticospinal excitability, hemiplegic cerebral palsy, motor lateralization, motor fibers integrity

### Supplementary Note 1

**Lateralization index for time-frequency regions of interest.** Baseline normalized power (ERS/ERD) was averaged across all time-frequency points of each time-frequency window, independently for contralateral (C) and ipsilateral (I) primary motor cortex of the moving hand. Lateralization index (LI) was then calculated for each window. For synchronization windows (theta ERS, beta rebound, gamma ERS), LI = (C – max(I, 0)) / (C + max(I, 0)). For desynchronization windows (mu ERD, beta ERD), LI = (C – min(I, 0)) / (C + min(I, 0)). The ipsilateral term was set to zero if its sign was opposite to the type of response in that window (negative for synchronization, positive for desynchronization). This way, an activity in the opposite direction could not be considered as the response in the window. For synchronization and desynchronization responses, LI is positive when contralateral activity is larger, negative when ipsilateral activity is larger, and equal to zero if both responses are equally strong.

### Supplementary Table 1

**Supplementary Table 1 Predictor weights, loadings, variable importance and collinearity for the partial least-squares structural-functional integrity score**

| Metric | weight | loading | VIP | VIF |
|---|---|---|---|---|
| DMF MD | -0.33 | -0.34 | 1.45 | 187.42 |
| DMF RD | -0.32 | -0.33 | 1.43 | 155.48 |
| Lesion size | -0.31 | -0.27 | 1.40 | 2.72 |
| MCF RD | -0.30 | -0.34 | 1.34 | 238.92 |
| DMF FA | 0.30 | 0.29 | 1.33 | 18.74 |
| MCF FA | 0.29 | 0.32 | 1.31 | 20.42 |
| MCF MD | -0.29 | -0.33 | 1.29 | 469.71 |
| $ERD_{beta}$ | -0.27 | -0.21 | 1.22 | 4.46 |
| DMF AD | -0.25 | -0.27 | 1.12 | 10.18 |
| MCF AD | -0.24 | -0.29 | 1.09 | 44.21 |
| $ERD_{mu}$ | -0.24 | -0.18 | 1.08 | 3.13 |
| $LI_{MRR}$ | 0.23 | 0.22 | 1.02 | 1.93 |
| Mu desync $LI_{TF}$ | -0.13 | -0.13 | 0.59 | 1.84 |
| Beta sync $LI_{TF}$ | 0.12 | 0.08 | 0.54 | 2.93 |
| Gamma sync $LI_{TF}$ | 0.06 | 0.08 | 0.28 | 2.60 |
| $ERS_{beta}$ | -0.03 | 0.01 | 0.14 | 2.45 |
| $ERS_{theta}$ | 0.02 | 0.07 | 0.11 | 2.88 |
| $ERS_{gamma}$ | -0.02 | 0.02 | 0.09 | 2.81 |
| Beta desync $LI_{TF}$ | -0.02 | -0.03 | 0.09 | 2.93 |
| Theta sync $LI_{TF}$ | 0.00 | -0.03 | 0.01 | 1.96 |

AD = axial diffusivity; DMF = descending motor fibers; ERD = event-related desynchronization; ERS = event-related synchronization; FA = fractional anisotropy; LI = lateralization index; MCF = motor commissural fibers; MD = mean diffusivity; MRR = movement-related response; RD = radial diffusivity; TF = time-frequency; VIF = Variance inflation factor; VIP = Variable importance in projection.

#### Supplementary Fig. 1

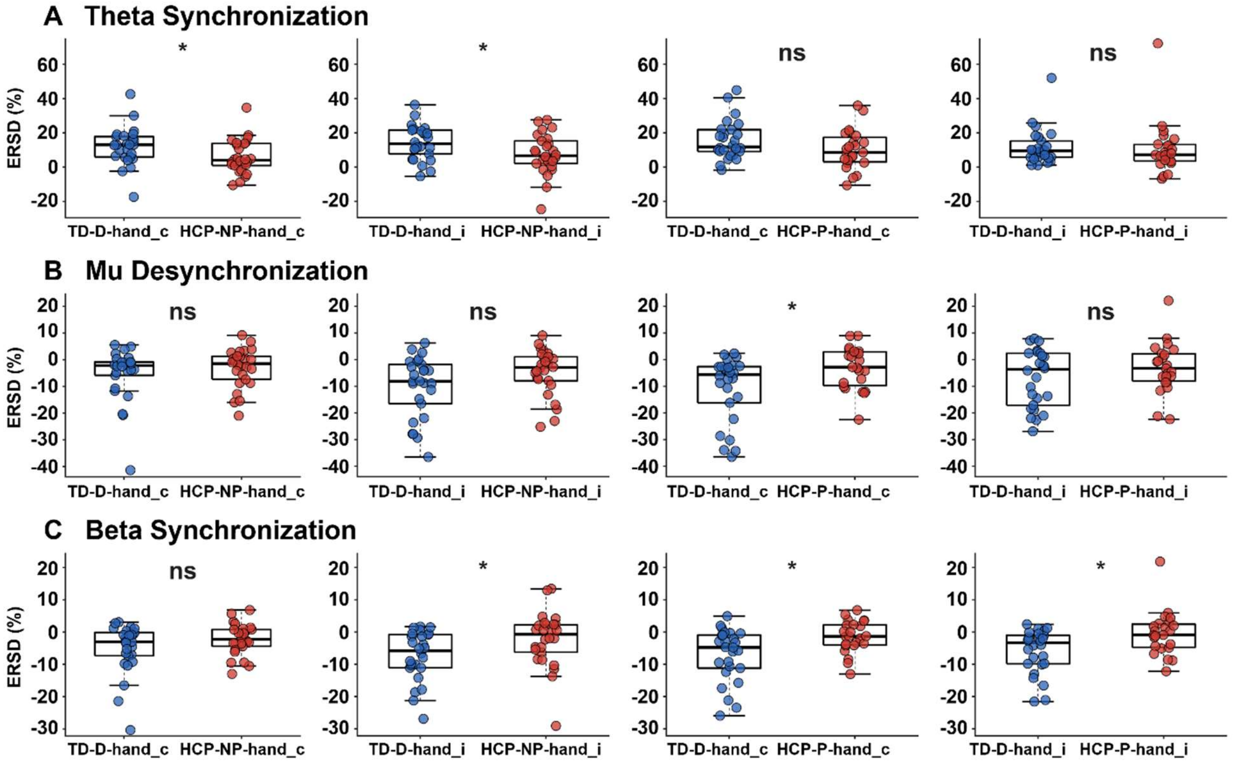


**Supplementary Fig. 1. Movement-related spectrotemporal activity**

Box-and-whisker plots of averaged movement-related **(A)** theta synchronization ($ERS_{theta}$), **(B)** mu desynchronization ($ERD_{mu}$), and **(C)** beta desynchronization ($ERD_{beta}$) during dominant-hand (D-hand) and non-dominant-hand (ND-hand) movements in typically developing (TD) controls as well as paretic hand (P-hand) and non-paretic hand (NP-hand) movements in children with hemiplegic cerebral palsy (HCP). Comparisons are shown for the contralateral (c) and ipsilateral (i) hemispheres during hand movements. Boxes indicate median and interquartile range, whiskers range, × denotes mean, and o represents individual data points for each participant. * $p < 0.05$, derived from Wilcoxon signed rank tests for within-subject and Mann-Whitney U tests for between-subject comparisons (FDR corrected for multiple comparisons). ERSD, event-related synchronization/desynchronization ;ns, non-significant.

### Supplementary Fig. 2

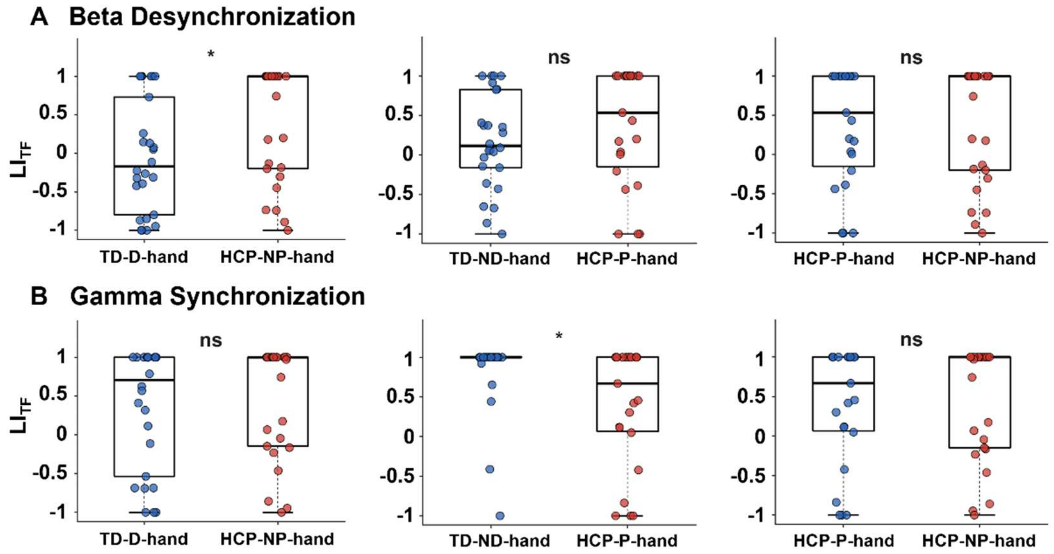


**Supplementary Fig. 2. Movement-related spectrotemporal motor lateralization**

Box-and-whisker plots of **(A)** beta desynchronization ($ERD_{beta}$) lateralization and **(B)** gamma synchronization ($ERS_{gamma}$) lateralization during unilateral finger movements in typically developing (TD) controls and children with hemiplegic cerebral palsy (HCP). Comparisons are shown between the TD dominant-hand (TD-D-hand) and HCP non-paretic-hand (HCP-NP-hand), TD non-dominant-hand (TD-ND-hand) and HCP paretic-hand (CP-P-hand), and the P-hand and NP-hand conditions within the HCP group. Time-frequency lateralization indices ($LI_{TF}$) quantify the relative contribution of contralateral and ipsilateral hemispheres. Boxes indicate median and interquartile range, whiskers range, × denotes mean, and o represents individual data points for each participant. *$p<0.05$, derived from Wilcoxon signed rank tests for within-subject and Mann-Whitney U tests for between-subject comparisons (FDR corrected for multiple comparisons). ns: non-significant.

#### Supplementary Fig. 3

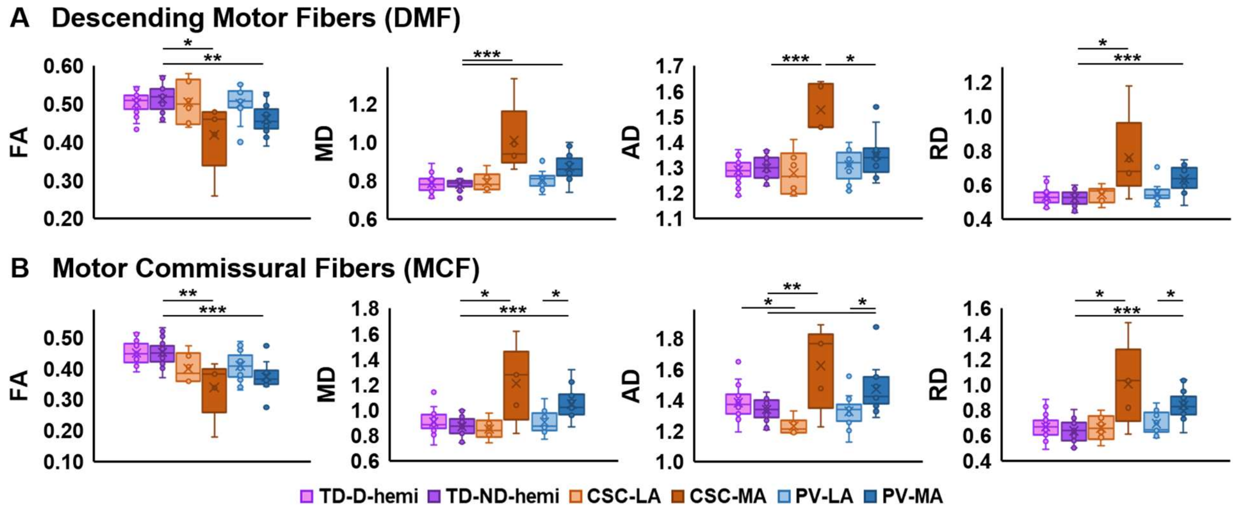


**Supplementary Fig. 3 Lesion-specific diffusivity metrics in motor white matter tracts**
Box-and-whisker plots of fractional anisotropy (FA), mean diffusivity (MD), axial diffusivity (AD), and radial diffusivity (RD) obtained from **(A)** descending motor fibers (DMF) and **(B)** motor commissural fibers (MCF) in the dominant (D-hemi) and non-dominant (ND-hemi) hemispheres of the typically developing (TD; N = 23) controls and the less affected (LA; N = 6) and more affected (MA, N = 5) hemispheres of the children with cortical-subcortical (CSC) and periventricular (PV; N = 14) lesions. Boxes indicate median and interquartile range, whiskers range, × denotes mean, and o represents individual data points for each participant. Diffusion metrics include fractional anisotropy (FA), mean diffusivity (MD), axial diffusivity (AD), and radial diffusivity (RD). *$p<0.05$, **$p<0.01$, ***$p<0.001$ derived from Wilcoxon signed rank tests for within subject, and Mann-Whitney U tests for between subject comparisons (FDR corrected for multiple comparisons).

#### Supplementary Fig. 4

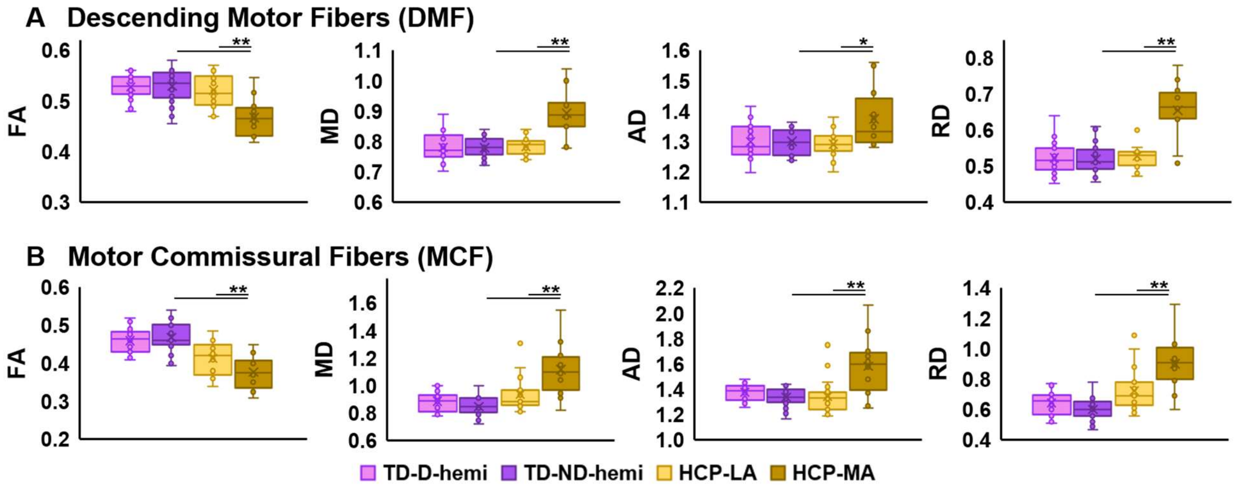


**Supplementary Fig. 4. Diffusivity metrics of motor white matter tracts reconstructed from nTMS-defined motor cortex**

Box-and-whisker plots of fractional anisotropy (FA), mean diffusivity (MD), axial diffusivity (AD), and radial diffusivity (RD) obtained from **(A)** descending motor fibers (DMF) and **(B)** motor commissural fibers (MCF) reconstructed using nTMS-defined primary motor cortex in the dominant (D-hemi) and non-dominant (ND-hemi) hemispheres of typically developing (TD) controls and the less affected (LA) and more affected (MA) hemispheres of children with hemiplegic cerebral palsy (HCP). Boxes indicate median and interquartile range, whiskers range, × denotes mean, and o represents individual data points for each participant. *$p<0.05$, **$p<0.01$, ***$p<0.001$ derived from Wilcoxon signed rank tests for within-subject and Mann-Whitney U tests for between-subject comparisons (FDR corrected for multiple comparisons).
